# Zoonotic and environmental sources of infant enteric pathogen infections identified with longitudinal sampling

**DOI:** 10.1101/2024.12.03.24318441

**Authors:** Abigail P. Paulos, John Mboya, Jeremy Lowe, Daehyun Daniel Kim, Hannah C. Wharton, Faith Thuita, Valerie L. Flax, Sammy M. Njenga, Angela R. Harris, Amy J. Pickering

## Abstract

Enteric pathogen infections in young children can result in diarrhea, malnutrition, and developmental impairments. Many enteric pathogens that infect young children can be zoonotic, yet the exposure risk of domestic animals living in close proximity to young children is poorly understood. We conducted a prospective cohort study with longitudinal microbiological sampling of child stool, animal feces, and the household environment to investigate pathogen transmission between animals and children under two years of age in pastoralist communities in rural Northern Kenya. We generated 28,743 pathogen-sample observations by measuring 33 bacterial, viral, protozoan, and helminth pathogens in the following types of samples (n=871) collected from households at four consecutive visits: child stool, caregiver stool, drinking water, food, child hands, household soil, and feces from ruminant, avian, and canine domesticated animals. Child enteric pathogen burden increased with age from a median of 1 pathogen among children under 3 months to 5 pathogens at 1-2 years old. Of the 20 unique pathogens detected in child stool, 85% were also detected in animal feces. New infections in children were associated with preceding household detection of the same pathogen in soil (Odds ratio: 8.8, 95% confidence interval: 3.3 – 23) and on child hands (Odds ratio: 5.0, 95% confidence interval 1.1 – 17). Network analysis revealed transmission of pathogens from poultry, dog, and ruminant feces to household soil, and between child hands and child stool. Our results indicate child hand contact with soil containing animal feces is a primary transmission route for first infections among children living in close proximity to animals. Our study provides new evidence that domestic animals in the household environment contribute to early-life enteric pathogen exposure, and that child hand hygiene could substantially prevent animal-human enteric pathogen exchange.

## Introduction

The annual burden of diarrhea in Africa is estimated at 1 billion cases and >500,000 deaths, with the majority of deaths occurring in children under 2 years old.^1^ The etiology of child diarrhea in Africa is diverse, including viruses (rotavirus, norovirus, adenovirus), bacteria (*Escherichia coli* pathotypes, *Campylobacter* spp.*, Shigella*), and protozoa (*Giardia duodenalis, Cryptosporidium).*^1,2^ Diarrheal and other enteric pathogens (e.g. helminths) also contribute to malnutrition, increasing susceptibility to future enteric infections and related morbidity and mortality.^3–5^ Fecal-oral transmission routes for enteric pathogens in settings with limited sanitation and hygiene infrastructure are complex; previous studies have detected fecal contamination in drinking water, food, soil, on hands, flies, and surfaces.^6–8^ While increased fecal contamination in drinking water and on child hands have been shown to be associated with child diarrhea, there have been limited studies that have measured contamination in other environmental reservoirs.^8^

Previous studies investigating environmental exposure pathways have mostly measured fecal indicator bacteria concentrations instead of actual pathogens. These studies often use a quantitative microbial risk assessment framework to estimate probability of illness, which requires strong assumptions about how fecal indicator bacteria relate to illness risk.^6,9–16^ Fecal bacteria levels in environmental samples are not good predictors of the presence of specific pathogens or markers of fecal contamination,^17,18^ and pathogen transmission pathways may substantially differ from fecal indicator bacteria due to differences in the fecal source and environmental fate and transport. A lack of direct measurements of viral, bacterial, protozoan, and helminth pathogens in the household environment limits our ability to elucidate key transmission pathways to young children.^19^

Exposure to animal feces is hypothesized to play a role in household enteric pathogen transmission. Prior research has found that living in close proximity to animals is associated with increased exposure to fecal contamination and increased child diarrhea.^17,20–23^ Host-specific fecal markers from animals (dogs, birds, and ruminants) were detected in multiple household reservoirs (soil, on hands, and in stored drinking water) in rural and urban Bangladesh.^17,24^ In Ecuador, identical sequence types of *C. jejuni* and atypical enteropathogenic *E. coli* were found in both child stool and household animals.^25^ However, measurement of pathogens in animal feces has been limited to cross-sectional studies sampling a small number of animals at the community level (e.g. not paired with human or environmental samples from the same household).^19,26^ Sampling child stool and domesticated animal feces from the same households over time is needed to understand the role of animals in early child enteric infections.

## Methods

### Study design

We used a prospective cohort study design to conduct extensive enteric pathogen testing of child stool, caregiver stool, animal feces, child hands, soil, food, and water from the household environment. Our goal was to identify sources of new infections in children under 1 year of age and possible environmental transmission pathways. The study was conducted in rural, pastoralist communities in the sub counties of Turkana South and Samburu North in northern Kenya. Prior county-wide surveys in Turkana and Samburu have found low access to improved WASH, high poverty rates, high rates of animal ownership, and elevated levels of child diarrhea and acute malnutrition.^27,28^ We enrolled children under 2 years of age as prior work suggests that the first two years of life are vital to long-term child growth trajectories and cognitive development.^29–31^

In total, we enrolled 100 households with a child under 2 years of age, with approximately equal enrollment across four key development age groups to characterize pathogen infection profiles in these age ranges: 0 - <3, 3 - <6, 6 - <12, and 12 - <24 months (**Fig 1**).^32^ Target age groups were selected based on Environmental Protection Agency (EPA) child development stages^32^ and to allow for the detection of first exposures to pathogens for young children. Sixty children <12 months were enrolled into a longitudinal cohort and visited up to four times approximately one week apart (a typical incubation period for many enteric pathogens) to investigate transmission pathways for new infections in early life. Children <12 months were prioritized for longitudinal sampling to study how first infections occur in early life, a critical period of development and growth.

**Figure 1.**
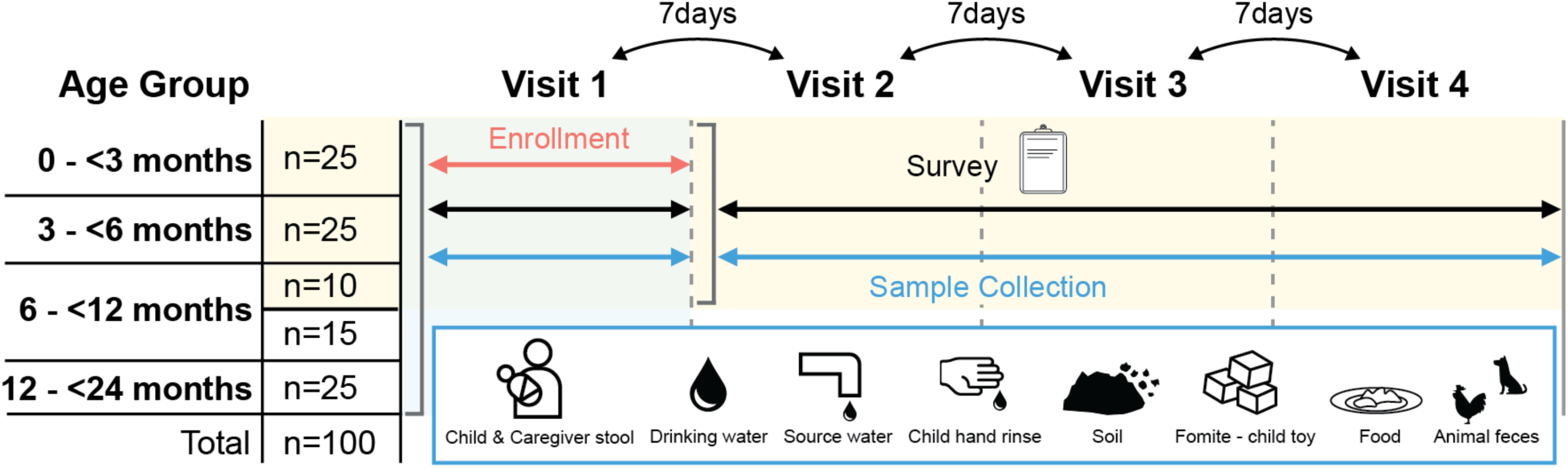
Conceptual diagram of study design. Twenty-five children were enrolled within each target age group (0 to <3, 3 to <6, 6 to <12, and 12 to <24 months). Children <6 months of age and a subset (n=10) of children between 6 and 12 months of age were sampled longitudinally across four total visits with 7 days between each visit; all other children >6 months were sampled once.

Within each sub county, we selected 10-15 villages for study based on travel feasibility (proximity to the field lab to enable processing within 6 hours) and field staff safety. In each village, we enrolled all households with a child under 2 years of age until we reached our target numbers in each child age group. In total, we enrolled 100 households (n=50 in each sub county). Approximately twenty-five households with a child in each target age group (0-2 months, 3-5 months, 6-11 months, and 12-23 months) were enrolled across both sub counties

In each household, we collected the following samples: child and caregiver stool, fresh animal feces, soil, stored food, stored drinking water, and child hand rinses. Samples were analyzed using a custom TaqMan Array Card (TAC) designed to detect 33 pathogens known to cause diarrhea or other child morbidity in Kenya or other LMICs (**Table S1**).

IRB approval for this study was obtained from North Carolina State University (24097) and Amref Health Africa in Kenya (P1055-2021), and an in-country research permit was obtained from NACOSTI.

### Study Site

This study was conducted between January and May 2022 in pastoralist and agro-pastoral communities in the Turkana South and Samburu North sub counties of arid and semi-arid lands regions of Kenya. The project was conducted in collaboration with the USAID Nawiri project in their program area, a USAID Bureau of Humanitarian Assistance (BHA) funded Development Food Security Activity designed to reduce persistent acute malnutrition in four counties of northern Kenya, including Turkana, Samburu, Marsabit and Isiolo. The primary occupation in the study areas is pastoralism and agro-pastoralism. Water insecurity in the area is exacerbated by droughts and flooding.^33^ For a description of access to water, sanitation, and hygiene (WASH) in the study setting, see the supporting information.

### Sample Collection

Stool samples were collected from the target child under the age of two years and each mother/caregiver in eligible households. Mothers were given two stool collection kits (for herself and her child) and instructed how to fill the containers. Field staff revisited the household the following morning and up to three times in total to collect the stool samples.

We collected fresh animal feces from one of each of the following types of animals when they were available in the household of the target child: chickens, cattle, goats, dogs, donkeys, sheep, and camels. The field staff observed animals defecating and collected feces as soon as possible after defecation; otherwise, our team asked respondents to point out where the freshest animal feces was located and the sample was collected from the ground. Veterinary technicians on the field team performed rectal palpation on goats, sheep, and cattle to obtain freshly defecated animal feces when possible, approximately 75% of animal fecal collection.

The soil surrounding the entrance to the household (<2 meters away from the entrance) was sampled as this location has been found to frequently contain high levels of fecal bacteria, as well as soil transmitted helminth eggs in Kenya and Tanzania during previous studies conducted by our team.^6,24,34^ Field staff identified stored weaning food to be served to children <2 years in the household and asked the respondent to provide a small amount of food in the same manner they feed their children, including serving it on a plate/bowl using any common utensils. Food types included porridge, chapati, potatoes, cassava, beans, rice, and others.

For drinking water sample collection, field staff asked the respondent to provide a glass of water as if giving it to their < 2 year-old child. Water was poured directly from the glass/cup into a sterile Whirlpak bag. Field staff collected > 250 mL of drinking water at each household. For hand rinse samples, field staff asked the respondent to place the target child’s left hand into a sterile Whirlpak bag pre-filled with 250 mL of distilled water and collected the sample as previously described.^35^

### Sample processing and analysis

All samples were preserved on ice in cooler boxes and transported to the field lab to be processed within 6 hours of sample collection using the IDEXX most probable number (MPN) method with Colilert media and QuantiTray 2000s to detect *E. coli* and total coliform. A 100 mL aliquot of each drinking water and child hand rinse sample was processed by membrane filtration using a 0.45 micron HA filter pre-treated with 2 mL of 1.25M of magnesium chloride. Filters were preserved using 0.5 mL of Zymo DNA/RNA Shield. Aliquots of the fecal, food, and soil samples were prepared and consisted of 0.3 g of sample in 750 uL of Zymo DNA/RNA Shield.

The ZymoBIOMICS DNA/RNA Miniprep Kit (Zymo Research) was used to extract nucleic acids from all animal feces, human stool, and food samples. Lysis buffer volume was increased by 2.5x to 1200 uL to ensure complete lysis in all samples. For soil samples, we used the RNeasy PowerSoil Total RNA Kit with the DNA Elution Kit add-on (Qiagen) to co-extract RNA and DNA. For both kits, we eluted in 100 µL and stored extracts at −80°C until TAC analysis.

We designed a custom TaqMan Array Card (TAC; Fisher) containing enteric pathogen targets (**Table S5**). The TAC is a platform for quantitative real-time PCR that enables simultaneous detection of up to 48 targets per sample in one run. We selected enteric pathogen targets associated with diarrhea or other diseases of child health significance, previously detected in Kenya or similar settings. In brief, we used 2-step reverse transcription followed by TAC to detect and quantify pathogens in nucleic acid extracts. Because of low nucleic acid content, food, child hand rinse, and drinking water samples were subjected to preamplification using the same set of primers on the custom TAC card between RT and TAC analysis (see Supporting Information for details). Primers and probes for assays included on the custom TAC are listed in **Table S9**, standard curve efficiencies in **Table S10**, sample limits of detection in **Table** S11, and gBlock sequences used for the standard curve in **Table S12**.

### Statistical Analyses

Outcomes of interest in the household risk factor models were child carriage of (1) any pathogen, (2) *E. coli* pathotypes, (3) bacterial pathogens, (4) viral pathogens, (5) protozoa or helminths and (6) EAEC, (7) EPEC, (8) ETEC-LT, (9) adenovirus, (10) norovirus GII, (11) *G. duodenalis*, and (12) enterovirus. The individual pathogens included were the seven most commonly detected pathogens and were detected in >10% of child stool. Exposure variables of interest included animal ownership, child age and mobility, latrine access, proximity of primary water source, antibiotic use, child breast milk consumption, child food consumption, and disposal of animal feces in the courtyard. We used generalized linear models to calculate odds ratios for risk factors, with standard errors clustered at the household. All households were included in this analysis. Adjusted models controlled for child age. Analyses were conducted in R version 4.2.3.

New infections were estimate for children enrolled in the longitudinal cohort and were defined as (1) no carriage of the pathogen in the prior round and (2) carriage in the current round. Pathogen presence in environmental samples were lagged by one visit and we calculated risk ratios for new infection in children given pathogen presence 7 days prior in (1) animal feces, (2) mother stool, (3) soil, (4) drinking water, and (5) any environmental sample (soil, child hands, food, or water). Odds ratios were estimated using Fisher’s exact method with confidence intervals. Odds of pathogen codetection were calculated between all pairs of sample types using Fisher’s exact method for confidence interval estimation.

To explore overlap between environmental pathways, we also calculated odds ratios for 1) leading pathogen contamination, defined as the same pathogen present in another sample type at the prior visit, and 2) pathogen co-detection, defined as the same pathogen present in multiple sample types on the same visit. Using these odds ratios, we created an aggregated pathogen transmission network which displays the number of connections across the 20 pathogens with significant leading pathogen odds ratios (**Fig 5**). We also created a network graph for each pathogen representing pathogen overlap across sample types, where each node is a sample type and each edge indicates an odds ratio with p < 0.05. The open source tool graphviz was used to generate networks graphs of pathogen overlap.^36^ Each edge represented overlap, defined as an odds ratio with a p-value less than 0.05. The width of each edge represents the magnitude of the odds ratio with the width increasing with odds ratio. Nodes which had no connections with other nodes were excluded. Three categories of animal feces were considered: ruminants (cows, sheep, and goats), chickens, and dogs.

## Results

A total of 100 households were enrolled into the study, including 60 into the longitudinal cohort; detailed participant enrollment and sample collection numbers are listed in **Tables S13 and S15**. We generated 28,743 observations (33 pathogens measured in 871 samples). Most (80%) samples contained at least one pathogen. The most frequently detected pathogens across all sample types were *G. duodenalis* (35%), enteroaggregative *E. coli* (EAEC: 30%), enteropathogenic *E. coli* (EPEC: 29%), enterotoxigenic *E. coli* (ETEC-LT: 22%), and Shiga toxin-like *E. coli* (STEC: 19%) (**Fig 2, Table S1**). In child stool, the most common pathogens were EAEC (56%), enterovirus (47%), EPEC (36%), *G. duodenalis* (20%), adenovirus 40/41 (15%), ETEC-LT (15%), and norovirus GII (12%). Overall, 87% of child stool samples contained at least one pathogen. In the longitudinal cohort, all 56 children were infected at least once across all visits. Children overall were infected with a median of 4 pathogens, and pathogen carriage increased with child age (t-tests; see **Fig 2**): a median of 1 pathogen for 0-2 months, 2 pathogens for 3-5 months, 3 pathogens for 6-11 months, and 5 pathogens for 12-23 months. Of the 20 different pathogens detected in child stool, 17 of these were also detected in animal feces, 17 in soil, 10 in drinking water, 5 in food, 17 on child hands, and 16 in caregiver stool.

**Figure 2.**
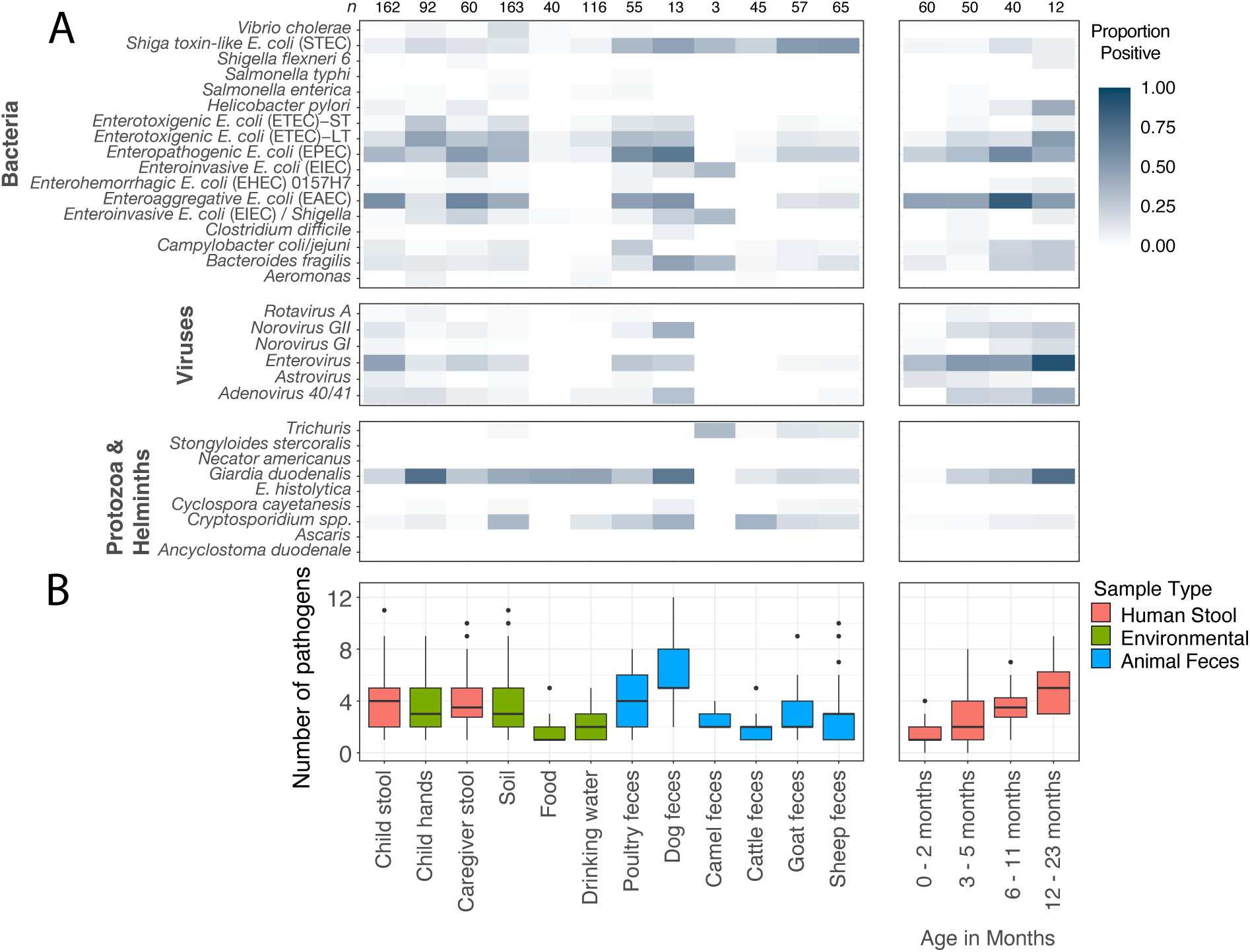
Enteric pathogen profiles across sample types: A) Proportion of samples positive for each pathogen across sample types (left) and in child stool by age (right). Sample sizes for each group are indicated at the top of the figure. B) Number of pathogens (out of a maximum of 33 pathogens) present by sample type and child age.

Animal feces carried a range of human enteric pathogens. To assess the diversity of pathogens in each sample type, we summed the number of different pathogens present in each sample out of a maximum of 33 pathogens (**Fig 2B**). Dog feces carried the greatest number of unique pathogens (median = 5); poultry feces had a median of 4 pathogens. Ruminant feces carried the smallest number of unique pathogens among all animal types (cattle: 2, goats: 2, and sheep: 3). Poultry and dogs were frequently infected with pathogenic *E. coli* (poultry: 76%, dogs: 92%; **Fig S1**), and infections of *G. duodenalis* (27%, 69%), enterovirus (27%, 23%), and non-parvum and non-hominis *Cryptosporidium* spp. (24%, 38%) were also common. Cattle also frequently carried STEC (22%), *Giardia* (11%) and *Cryptosporidium spp.* (38%). Goats and sheep were most commonly infected with STEC (goats: 51%, sheep: 53%), EPEC (23%, 23%), *Cryptosporidium* spp. (18%, 15%), and *G. duodenalis* (19%, 18%).

Food contained a median of 1 enteric pathogen while drinking water contained a median of 2 pathogens; for both sample types, the pathogen most commonly detected was *G. duodenalis* (food: 45%, water: 46%). Child hands carried a median of 3 pathogens per child, primarily *G. duodenalis* (74%) and forms of pathogenic *E. coli* (EAEC: 14%, EPEC: 24%; ETEC-LT: 46%; ETEC-ST: 27%); 77 of 92 child hand samples contained at least one pathogen. Soil contained a median of 3 pathogens, primarily *G. duodenalis* (42%), *Cryptosporidium spp.* (35%), and forms of pathogenic *E. coli* (EAEC: 42%, EPEC: 37%, ETEC-LT: 36%, ETEC-ST: 15%). For pathogen quantities by sample type, see Figs S2 – S6.

Approximately 75% of stored drinking water samples contained culturable *E. coli*, and over a third of all other sample types tested positive for *E. coli* by culture: 44% of food, 48% of child hands, 45% of fomites, 62% of soil. Pathogen concentrations in all samples were calculated with TAC results using a standard curve (**Fig 3; Table S10**). Across food, soil, drinking water, and child hands, we found few associations between pathogen and *E. coli* culture presence or concentrations using Spearman’s correlation tests: *E. coli* concentrations in culture were associated with EIEC on hands (r = 0.47, p = 0.05), ETEC-LT in drinking water (r = 0.56, p < 0.01), and norovirus GII in soil (r = 0.42, p = 0.01) and on hands (r = 0.47, p = 0.05; **Table S8**). *E. coli* concentrations measured by TAC (*uidA* gene) were associated with *E. histolytica* on hands (r = 0.68, p < 0.01) and in drinking water (r = 1.00, p < 0.01), astrovirus in chicken feces (r = 0.78, p < 0.01), and EPEC in goat feces (r = 0.71, p < 0.01; **Table S7**). For sample numbers processed for *E. coli* culture, see **Table S15**.

**Figure 3.**
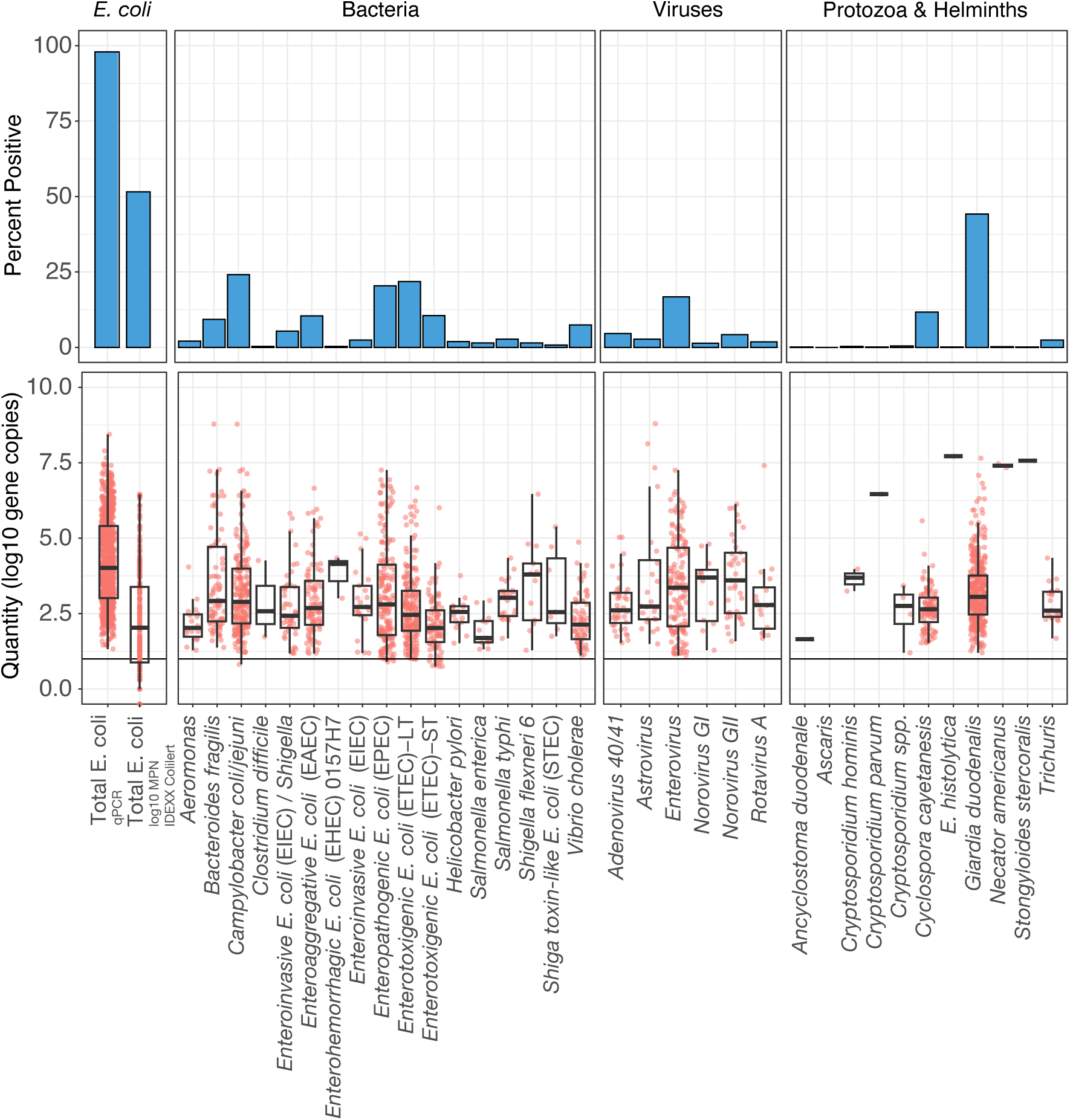
Sample positivity by pathogen (top) and distribution of quantities among positive samples measured by TAC by pathogen (bottom) (n=732 samples). The *E. coli* category includes the quantity of total *E. coli* detected by both qPCR and MPN by culture.

### Longitudinal analysis of environmental pathways for new infections

In total, we detected 33 new infections across 10 of the 33 measured pathogens, with new infections detected in 20 of the 56 children (36%) sampled longitudinally (4 children were unavailable for follow up visits). Children with new infections had a median of 1 new infection; 7 of 20 children were newly infected with more than one pathogen in the same visit and 3 of 20 children were newly infected across multiple sampling rounds. The number of new infections decreased with child age; we observed 15 in children 0 – 2 months (73% of children in this age group), 12 in 3 – 5 months (60%), and 6 in 6 – 11 months (50%). Children were most often newly infected with EPEC (n = 8), EAEC (8), enterovirus (5), norovirus GII (4), *G. duodenalis* (2), and adenovirus (2). Pathogens responsible for 1 new infection included STEC, ETEC-LT, astrovirus, and *Campylobacter coli/jejuni.* Overall, across the 33 new infections, we identified the same pathogen in any type of pathway (environmental samples, animal feces, or mother stool) on the prior visit in 16 (48%) cases.

Pathogen detection in soil in the prior sampling round was associated with new infections (Odds ratio [OR]: 8.8, 95% CI: 3.3 – 23, p < 0.01; **Fig 4**), as was pathogen detection on child hands (OR:5.0, 95% CI: 1.1 – 17, p = 0.02). When aggregated across all environmental pathways (hands, soil, water, and food, when available), we find that pathogen detection in environmental pathways was associated with new infections (OR: 6.1, 95% CI: 2.7 – 14, p < 0.01). We aggregated results of all animal feces types (due to limited power to investigate by specific animal host), yet we found no statistically significant association between new infections and pathogen presence in animal feces on the previous visit (OR: 2.2 95% CI: 0.6 – 6.3, p=0.15). We found no associations between new infections in child feces and food, water samples, or mother stool as there was limited overlap in the specific pathogens found in child stool and these sample types; the same pathogen which newly infected a child was detected in 0 food samples, 2 water samples, and 2 mother stool samples 7 days prior. We also assessed the odds of child infection given pathogen detection in each sample type either on the prior visit or the same visit; we find that pathogen detection in animal feces and mother stool are associated with new infections (**Fig S7**).

**Figure 4.**
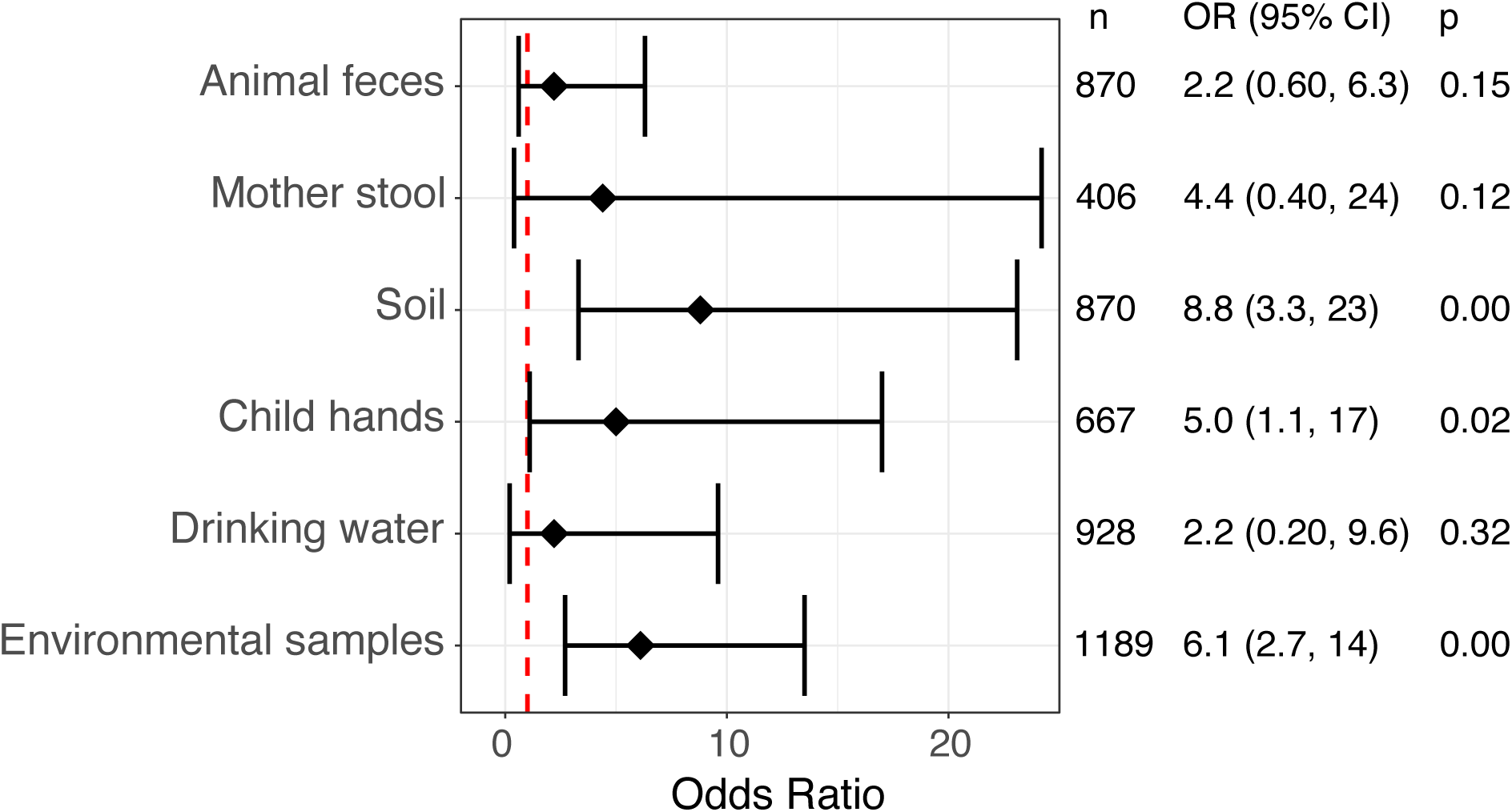
Odds ratios of pathogen detection in animal feces or environmental samples and subsequent new infection with the same pathogen in children. Bars represent 95% confidence intervals. The dotted red line indicates an odds ratio of 1. The environmental samples category includes hands, soil, water, and food. Food alone is not displayed because of large uncertainty around the point estimate.

Food sample collection in the household was limited as few households had food available at the time of collection, or caregivers solely breastfed their child; only n=40 total samples were collected, compared to n=116 for drinking water and n=163 for soil. However, we estimated the odds of pathogen detection in food given detection in soil or animal feces, and we find that the odds of pathogen detection in food was 13 times higher when that pathogen was present in either soil or animal feces on the same visit (OR: 13, 95% CI: 7.5, 23).

### Pathogen transmission networks

Across 13 of 20 pathogen networks, there was a connection between child hands and child stool, highlighting the role that hands play in transmission of these pathogens (**Fig 5, Fig S8**). Of the pathogen networks with an exposure associated with subsequent child stool infection, hands were the most common (6 of 10), followed by soil (4 of 10), chicken feces (4 of 10), caregiver stool (1 of 10), dog feces (1 of 10), and food (1 of 10). Animal feces had a connection to soil in 9 of 20 pathogen transmission networks, and a connection to any other environmental pathway in 16 of 20 networks. Detection in ruminant and chicken feces preceded caregiver infection for *Campylobacter,* sample types for 16 of 20 pathogens. For thirteen pathogens, no significant relationships were found between any sample types due to rare or no detections.

**Figure 5.**
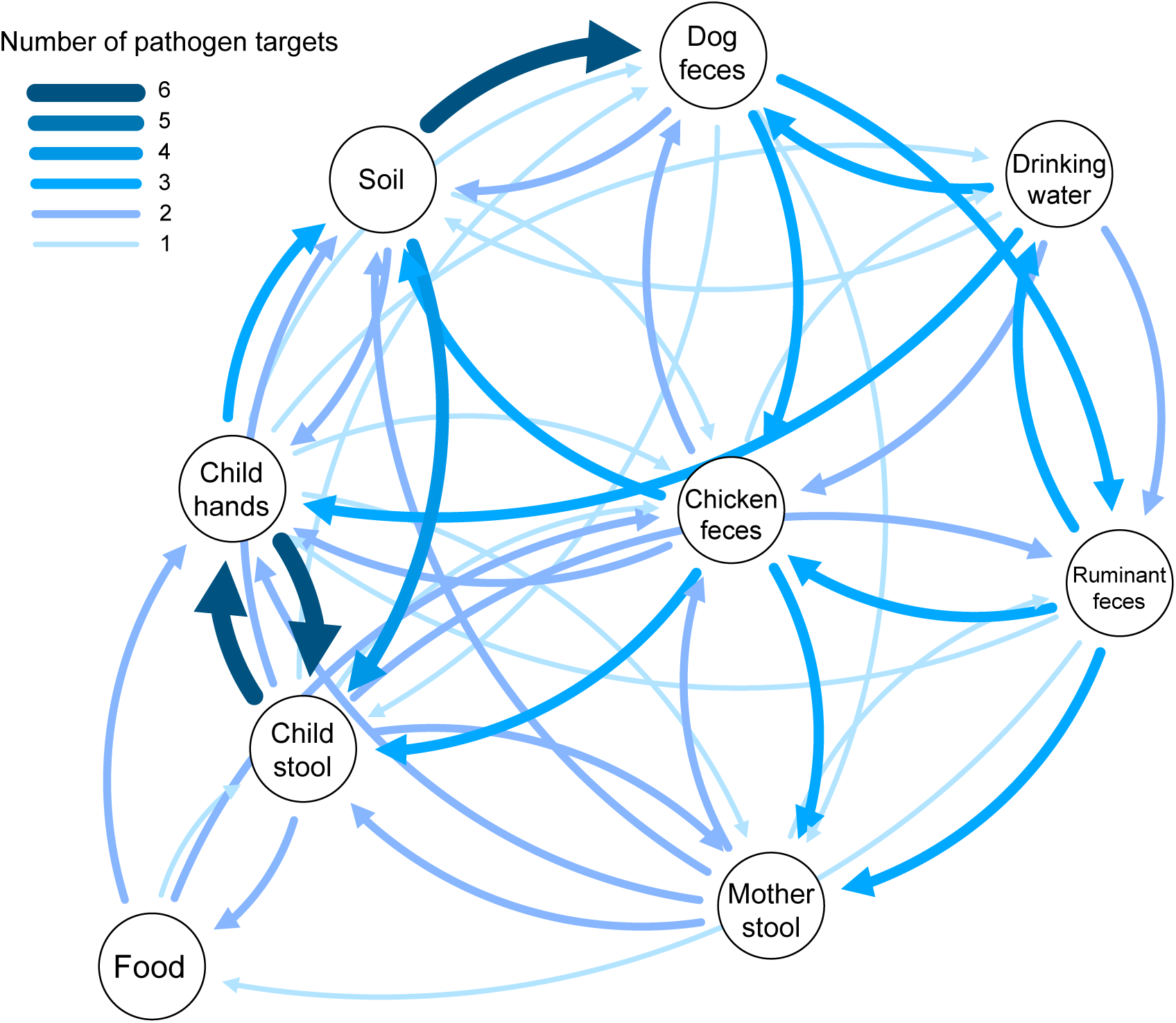
Pathogen transmission network graph. Nodes represent the sample type. Edges represent the number of pathogens for which detection in the source node was associated with detection in the receiving node on the next visit within the same household; edge thickness and color correspond to the number of pathogen targets with a significant (p < 0.05) relationship.

### Household characteristics associated with child pathogen carriage

To understand how household factors relate to child infection status, we calculated odds ratios of child pathogen carriage for risk factors recorded in our household surveys. Risk factors considered included those related to child behavior, including mobility (crawls or walks) and diet (receiving breast milk or food), as well as household characteristics such as latrine access, livestock ownership, food access, and disposal of animal feces in the courtyard. Outcomes considered included carriage of any pathogen, viral pathogens, bacterial pathogens, *E. coli* pathotypes, protozoa/helminths, EAEC, ETEC-LT, EPEC, adenovirus, enterovirus, *G. duodenalis*, and norovirus GII. The seven pathogens included as outcomes in separate models were the 7 most commonly detected pathogens.

Significant risk factors varied by pathogen type (**Table 1**). Child consumption of food as compared to exclusive breastfeeding was associated with increased odds of detection of protozoa/helminths, EAEC, adenovirus, enterovirus, and *G. duodenalis* (**Tables S2-S6**). Falling below the poverty line was associated with increased carriage of pathogens overall and of pathogenic *E. coli* and bacteria pathogens. Broadly, infection risk of all types increased with child age and child mobility: crawling was associated with carriage of protozoa/helminths and *G. duodenalis*, and walking with EAEC, EPEC, ETEC, and norovirus GII. For bacterial pathogens overall, EAEC, and ETEC-LT, mother usage of antibiotics during pregnancy was associated with increased odds of child pathogen carriage. Dog ownership was associated with increased odds of norovirus GII carriage, and cattle ownership with enterovirus and norovirus GII carriage. Long walk time to the primary water source for the household was associated with increased odds of bacterial pathogens and norovirus GII carriage. Latrine ownership was protective against EPEC infections.

**Table 1.**
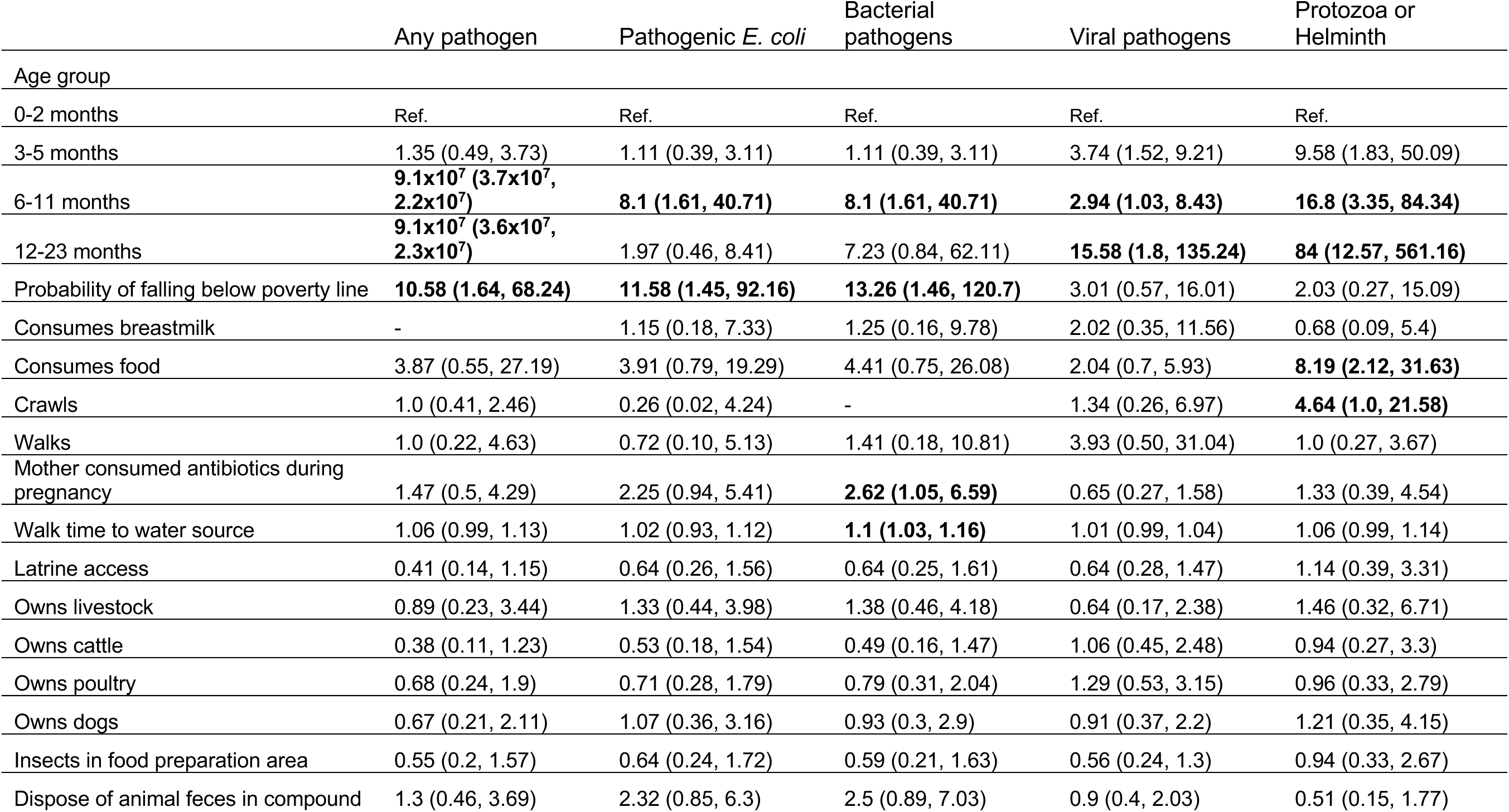
Household risk factors for child enteric pathogen infections. Odds ratios and 95% confidence intervals shown, adjusted for child age group. Bold indicates p<0.05.

Although not a main objective of this study because of the limited number of children enrolled with diarrhea, we find that rotavirus was significantly and positively associated with caregiver-reported child diarrhea at the time of sample collection (**Table S16**); for diarrheal prevalences in the study, see **Table S14**).

## Discussion

Our results provide the first longitudinal evidence that animal feces are an important source of initial and early life (<12 months) child enteric infections. We found substantial overlap of pathogens in child stool and animal feces; 85% of pathogens detected in child stool were also detected in animal feces. Detection of a pathogen in animal feces within a household was significantly associated with child infection with the same pathogen (**Fig S7**). Child hands and soil were important environmental transmission routes for infant infections, particularly for pathogens originating in chicken feces (**Fig 5)**. Detection of Adenovirus, Astrovirus, *Cryptosporidium*, *G. duodenalis,* and several *E. coli* pathotypes on a child’s hands frequently preceded child infection with these pathogens. Soil contamination with Astrovirus, Norovirus, and *E. coli* pathotypes often preceded child infection. These findings highlight that hands and soil are transmission pathways for child-animal pathogen exchange within the domestic environment. As livestock are a critical source of household income and nutrition for young children,^37^ our results indicate a need for improved community and household infrastructure and practices to separate children from animal feces.

This study generated new evidence of animal fecal shedding of a wide range of human enteric pathogens. Some of these pathogens, such as pathogenic *E. coli*, *Campylobacter* spp., and *G. duodenalis*, are known or suspected to be zoonotic, while others such as enterovirus, adenovirus, and norovirus are not typically considered zoonotic.^21^ We also demonstrate that animal feces were widespread in the household environment, even in households that did not report owning animals (see SI text result section on household characteristics). Human feces is one potential initial source of these pathogens in animals, as poor human waste management in the study area may enable animal consumption of human feces and result in reverse zoonotic transmission (transmission from humans to animals).^38^ Notably, we find that detection of pathogens in child stool is associated with subsequent contamination of soil with the same pathogen, suggesting that poor management of child waste could also contribute to pathogen proliferation in the environment and, potentially, animal infections. Provision of safely managed sanitation could prevent reverse zoonotic transmission, yet prior work has found that provision of basic sanitation alone is not adequate to reduce enteric pathogen infection prevalence in children under 5 years.^39^ Safely managed sanitation infrastructure that integrates both animal feces and child feces management may be necessary to prevent zoonotic transmission.

The household environment in our study site was widely contaminated with *Giardia duodenalis* across all sample types. *G. duodenalis* is a leading cause of diarrhea in Africa, with an estimated 36,116 annual cases per 100,000 people.^1^ The prevalence of *G. duodenalis* infections in children in our study (20%) exceeded that of Kenya overall (9%),^40^ but was lower than the prevalence found in healthy children in other settings.^41^ Prior to this study, contaminated food and drinking water were well known transmission pathways for *G. duodenalis*.^42^ Here, we observed animal feces contaminating household soil and child hands with *G. duodenalis*. Nearly half of all soil samples contained *G. duodenalis*, compared to 18% of samples in an urban community in Kisumu, Kenya.^43^ Contamination with *G. duodenalis* on child hands (74%) was much higher than has previously been reported; a meta-analysis found that less than 2% of mother hands carried *G. duodenalis*.^17,44^ *G. duodenalis* has been previously detected in the feces of both livestock and household pets, which aligns with our observation of *G. duodenalis* in >50% of dog and >25% poultry feces.^21^ Notably, the prevalence of *G. duodenalis* in dog feces in our rural study site was approximately 3 times higher compared to dogs in an urban community in Kenya.^45^ Detection of *G. duodenalis* on child hands and in dog feces temporally preceded child infection; it follows that we identified crawling (involving frequent hand to soil contact) as a risk factor for child *G. duodenalis* infection. Having a dirt floor and chicken ownership were both risk factors for *G. duodenalis* infections in children under 2 in previous studies.^46,47^ Our findings indicate animal feces as an important source of child *G. duodenalis* infections through child hand contact with soil in rural, low-income settings.

Many prior studies have used *E. coli* as an indicator of fecal contamination in environmental reservoirs, and studies measuring bacterial, viral, protozoan, and helminth enteric pathogens have been limited.^19,48^ However, we found that *E. coli* is generally not associated with enteric pathogen detection in soil, food, drinking water, or on child hands. Of the most commonly detected pathogens in this study (EPEC, EAEC, ETEC, STEC, enterovirus, *G. duodenalis*, adenovirus 40/41, and norovirus GII), *E. coli* detection in an environmental pathway was only associated with detection of ETEC-LT in drinking water and norovirus GII in soil and hands. Accordingly, *E. coli* detection in these environmental reservoirs does not imply the presence of any specific enteric pathogens. Published studies conducting quantitative microbial risk assessments (QMRAs) often infer specific pathogen quantities in environmental reservoirs by assuming a ratio between measured *E. coli* and other pathogens.^49^ Our results suggest that this approach will not provide valid estimates of enteric pathogen levels, as there was no association between *E. coli* levels and most pathogens across multiple environmental media. Other studies using TAC have also found variable pathogen detection despite *E. coli* presence, as in Capone *et al*.^48^

Reported diarrheal rates in our study were high, and the pathogens we detected among children suggest different diarrhea etiologies in this setting than reported in prior studies. In the Global Enteric Multicenter Study (GEMS), the pathogens associated with diarrhea in children <12 months in Nyanza Province, Kenya were found to be rotavirus (22% of cases), *Cryptosporidium* spp. (9.9%), ETEC (8.8%), and adenovirus 40/41 (8.5%). For children between 12 and 24 months of age, the most important pathogens were EIEC/*Shigella* (15.4%), rotavirus (14.5%), ETEC (9.5%), and *Cryptosporidium* spp. (6.3%).^2^ A recent meta-analysis found that in East Africa, the pathogens responsible for the greatest diarrheal cases in children under 5 were EAEC, *G. duodenalis*, ETEC, and norovirus, while those responsible for the greatest number of deaths were EIEC, rotavirus, and *Cryptosporidium* spp.^1^ In this study, the prevalence of these pathogens in child stool was low, with the exception of EAEC, *G. duodenalis* adenovirus 40/41; however, diarrheal prevalence was high, and we found that child diarrhea was associated with rotavirus. Notably, GEMS was located in a region with different climate, culture, and animal husbandry practices than our study site, all of which may impact prevailing pathogen profiles. Prior research has found that practices of nomadic and pastoral peoples can increase the risk of zoonotic enteric parasite transmission;^50^ we hypothesize that these practices may also influence the types of enteric pathogens present and the key pathways for transmission.

Diarrhea has been associated with an increased risk of acute malnutrition in young children,^51–53^ as previously demonstrated in the USAID Nawiri longitudinal study in Samburu and Turkana Counties.^54,55^ The high prevalence of diarrhea in this study (>30% 7-day prevalence), and particularly the association we find between diarrhea and rotavirus, the most common cause of fatal diarrhea in children,^56^ could be contributing to the high prevalence of wasting in these two counties. Future research examining how early-life enteric pathogen exposure and infection contributes to wasting would be valuable to determine the potential for interventions that prevent pathogen transmission to reduce wasting.

This study has several limitations. Sample collection occurred during the dry season, and pathogen transmission pathways could be different during the rainy season and periods of heavy precipitation. The incubation time from initial exposure to symptomatic illness and/or pathogen shedding in child feces varies by pathogen, but sample collection was standardized to occur 1-2 weeks apart to capture most incubation periods. While qPCR assays have the advantage of uniquely identifying pathogens, they are unable to identify if the exact same strain is present across samples, as whole genome analysis can. However, this study is the first to apply longitudinal monitoring of a wide range of pathogens by qPCR across child and caretaker stool, animal feces, and investigate a comprehensive set of environmental transmission pathways.

Here, we demonstrated that children become infected with pathogens shortly after birth, despite reported exclusive breastfeeding. In this setting, the youngest children (< 6 months of age) quickly became infected with *E. coli* pathotypes, enterovirus, and *G. duodenalis.* Children under 6 months of age, and especially children under 3 months, are often excluded from studies of pathogen infection.^12,57^ We detected significant pathogen overlap between child hands and child stool, suggesting that hand contamination often precedes child infection. Other studies have reported that hand-to-mouth contacts can be an important transmission pathway for children over 3 months of age,^9,14,18,58,59^ and we extend these findings to those under 3 months of age. Typically, community hand hygiene campaigns focus on caregiver hand hygiene while young (<2 years) child hand hygiene is often left out of behavior change messaging. Altogether with previous evidence, our findings suggest that infant hand hygiene could substantially prevent early life pathogen infections, including zoonotic pathogens.

## Code and Data Availability

The raw data (de-identified) and code used for analyses are available at https://github.com/abharv52/infant_enteric_pathogen_transmission.

## Data Availability

All data produced in the present study are available upon reasonable request to the authors

## Acknowledgements

This work was funded by a grant to the Innovations for Poverty Action, Kenya from the PRO-WASH program funded by the United States Agency for International Development. AJP is a Chan Zuckerburg Biohub San Francisco Investigator.

## Notes

### Competing Interest Statement

The authors have declared no competing interest.

### Funding Statement

USAID PRO-WASH program

### Summary of Updates

We have updated the abstract and presentation of results.

## References

1. Thystrup, C. et al. Etiology-specific incidence and mortality of diarrheal diseases in the African region: a systematic review and meta-analysis. BMC Public Health 24, 1864 (2024).

2. Liu, J. et al. Use of quantitative molecular diagnostic methods to identify causes of diarrhoea in children: a reanalysis of the GEMS case-control study. The Lancet 388, 1291–1301 (2016).

3. Tickell, K. D. et al. The effect of acute malnutrition on enteric pathogens, moderate-to-severe diarrhoea, and associated mortality in the Global Enteric Multicenter Study cohort: a post-hoc analysis. Lancet Glob. Health 8, e215–e224 (2020).

4. Brown, K. H. Diarrhea and Malnutrition. J. Nutr. 133, 328S–332S (2003).

5. Control, N. R. C. (US) S. on N. and D. D. NUTRITIONAL CONSEQUENCES OF ACUTE DIARRHEA. in Nutritional Management of Acute Diarrhea in Infants and Children (National Academies Press (US), 1985).

6. Pickering, A. J. et al. Fecal Indicator Bacteria along Multiple Environmental Transmission Pathways (Water, Hands, Food, Soil, Flies) and Subsequent Child Diarrhea in Rural Bangladesh. (2018).

7. Robb, K., et al. Assessment of Fecal Exposure Pathways in Low-Income Urban Neighborhoods in Accra, Ghana: Rationale, Design, Methods, and Key Findings of the SaniPath Study. Am. J. Trop. Med. Hyg. 97, 1020–1032 (2017).

8. Goddard, F. G. B. et al. Faecal contamination of the environment and child health: a systematic review and individual participant data meta-analysis. *Lancet Planet*. Health 4, e405–e415 (2020).

9. Mattioli, M. C. M., Davis, J. & Boehm, A. B. Hand-to-Mouth Contacts Result in Greater Ingestion of Feces than Dietary Water Consumption in Tanzania: A Quantitative Fecal Exposure Assessment Model. Environ. Sci. Technol. 49, 1912–1920 (2015).

10. Wang, Y., et al. Multipathway Quantitative Assessment of Exposure to Fecal Contamination for Young Children in Low-Income Urban Environments in Accra, Ghana: The SaniPath Analytical Approach. Am. J. Trop. Med. Hyg. 97, 1009–1019 (2017).

11. Kwong, L. H. et al. Ingestion of Fecal Bacteria along Multiple Pathways by Young Children in Rural Bangladesh Participating in a Cluster-Randomized Trial of Water, Sanitation, and Hygiene Interventions (WASH Benefits). Environ. Sci. Technol. 54, 13828–13838 (2020).

12. Kwong, L. H. et al. Age-related changes to environmental exposure: variation in the frequency that young children place hands and objects in their mouths. J. Expo. Sci. Environ. Epidemiol. 30, 205+ (2020).

13. Null, C. et al. Effects of water quality, sanitation, handwashing, and nutritional interventions on diarrhoea and child growth in rural Kenya: a cluster-randomised controlled trial. (2018).

14. Mattioli, M. C. et al. Enteric Pathogens in Stored Drinking Water and on Caregiver’s Hands in Tanzanian Households with and without Reported Cases of Child Diarrhea. PLoS ONE 9, 1–11 (2014).

15. Uprety, S. et al. Assessment of microbial risks by characterization of Escherichia coli presence to analyze the public health risks from poor water quality in Nepal. Int. J. Hyg. Environ. Health 226, 113484 (2020).

16. Gruber, J. S., Ercumen, A. & Jr, J. M. C. Coliform Bacteria as Indicators of Diarrheal Risk in Household Drinking Water: Systematic Review and Meta-Analysis. PLOS ONE 9, e107429 (2014).

17. Fuhrmeister, E. R. et al. Predictors of Enteric Pathogens in the Domestic Environment from Human and Animal Sources in Rural Bangladesh. Environ. Sci. Technol. 53, 10023–10033 (2019).

18. Mattioli, M. C., Pickering, A. J., Gilsdorf, R. J., Davis, J. & Boehm, A. B. Hands and Water as Vectors of Diarrheal Pathogens in Bagamoyo, Tanzania. Environ. Sci. Technol. 47, 355–363 (2013).

19. Lappan, R. et al. Monitoring of diverse enteric pathogens across environmental and host reservoirs with TaqMan array cards and standard qPCR: a methodological comparison study. *Lancet Planet*. Health 5, e297–e308 (2021).

20. Ercumen, A. et al. Animal Feces Contribute to Domestic Fecal Contamination: Evidence from E. coli Measured in Water, Hands, Food, Flies, and Soil in Bangladesh. Environ. Sci. Technol. 51, 8725–8734 (2017).

21. Delahoy, M. J. et al. Pathogens transmitted in animal feces in low- and middle-income countries. Int. J. Hyg. Environ. Health 221, 661–676 (2018).

22. Barnes, A. N., Anderson, J. D., Mumma, J., Mahmud, Z. H. & Cumming, O. The association between domestic animal presence and ownership and household drinking water contamination among peri-urban communities of Kisumu, Kenya. PLOS ONE 13, e0197587 (2018).

23. Ercumen, A. et al. Poultry Ownership Associated with Increased Risk of Child Diarrhea: Cross-Sectional Evidence from Uganda. Am. J. Trop. Med. Hyg. 102, 526–533 (2020).

24. Boehm, A. B. et al. Occurrence of Host-Associated Fecal Markers on Child Hands, Household Soil, and Drinking Water in Rural Bangladeshi Households. Environ. Sci. Technol. Lett. 3, 393–398 (2016).

25. Vasco, K., Graham, J. P. & Trueba, G. Detection of Zoonotic Enteropathogens in Children and Domestic Animals in a Semirural Community in Ecuador. Appl. Environ. Microbiol. 82, 4218–4224 (2016).

26. Penakalapati, G. et al. Exposure to Animal Feces and Human Health: A Systematic Review and Proposed Research Priorities. Environ. Sci. Technol. 51, 11537–11552 (2017).

27. Turkana County SMART Nutrition Surveys. (2019).

28. Samburu County SMART Survey Report. (2019).

29. Black, R. E. et al. Maternal and child undernutrition and overweight in low-income and middle-income countries. The Lancet 382, 427–451 (2013).

30. Crookston, B. T. et al. Impact of early and concurrent stunting on cognition. Matern. Child. Nutr. 7, 397–409 (2011).

31. Georgiadis, A. & Penny, M. E. Child undernutrition: opportunities beyond the first 1000 days. Lancet Public Health 2, e399 (2017).

32. U.S. Environmental Protection Agency. Exposure Factors Handbook 2011 Ed.

33. Balfour, N., Obando, J. & Gohil, D. Dimensions of water insecurity in pastoralist households in Kenya. Waterlines 39, 24–43 (2020).

34. Pickering, A. J. et al. Fecal Contamination and Diarrheal Pathogens on Surfaces and in Soils among Tanzanian Households with and without Improved Sanitation. Environ. Sci. Technol. 46, 5736–5743 (2012).

35. Pickering, A. J., Boehm, A. B., Mwanjali, M. & Davis, J. Efficacy of Waterless Hand Hygiene Compared with Handwashing with Soap: A Field Study in Dar es Salaam, Tanzania. Am. J. Trop. Med. Hyg. 82, 270–278 (2010).

36. Gansner, E. R. & North, S. C. An open graph visualization system and its applications to software engineering. Softw. Pract. Exp. 30, 1203–1233 (2000).

37. Swarthout, J. M. et al. Addressing Fecal Contamination in Rural Kenyan Households: The Roles of Environmental Interventions and Animal Ownership. Environ. Sci. Technol. 58, 9500–9514 (2024).

38. Barnes, A. N. et al. Zoonotic enteric parasites in Mongolian people, animals, and the environment: Using One Health to address shared pathogens. PLoS Negl. Trop. Dis. 15, e0009543 (2021).

39. Fuhrmeister, E. R. et al. Effect of Sanitation Improvements on Pathogens and Microbial Source Tracking Markers in the Rural Bangladeshi Household Environment. Environ. Sci. Technol. 54, 4316–4326 (2020).

40. Tawana, M., Onyiche, T. E., Ramatla, T. & Thekisoe, O. A ‘One Health’ perspective of Africa-wide distribution and prevalence of Giardia species in humans, animals and waterbodies: a systematic review and meta-analysis. Parasitology 150, 769–780.

41. Alemu, Y. et al. Prevalence and assemblage of Giardia duodenalis in a case-control study of children under 5 years from Jimma, Southwest Ethiopia. Parasitol. Res. 123, 38 (2023).

42. Dixon, B. R. *Giardia duodenalis* in humans and animals – Transmission and disease. Res. Vet. Sci. 135, 283–289 (2021).

43. Baker, K. K. et al. Fecal Fingerprints of Enteric Pathogen Contamination in Public Environments of Kisumu, Kenya, Associated with Human Sanitation Conditions and Domestic Animals. Environ. Sci. Technol. 52, 10263–10274 (2018).

44. Mertens, A. et al. Effects of water, sanitation, and hygiene interventions on detection of enteropathogens and host-specific faecal markers in the environment: a systematic review and individual participant data meta-analysis. *Lancet Planet*. Health 7, e197–e208 (2023).

45. Maingi, H. N. Prevalence, Risk Factors and Geospatial Distribution of Giardia Duodenalis Infection in Dogs in Nairobi County, Kenya. (University of Nairobi, 2021).

46. Rogawski, E. T. et al. Determinants and Impact of Giardia Infection in the First 2 Years of Life in the MAL-ED Birth Cohort. J. Pediatr. Infect. Dis. Soc. 6, 153–160 (2017).

47. Benjamin-Chung, J. et al. Household finished flooring and soil-transmitted helminth and Giardia infections among children in rural Bangladesh and Kenya: a prospective cohort study. Lancet Glob. Health 9, e301–e308 (2021).

48. Capone, D., et al. Urban Onsite Sanitation Upgrades and Synanthropic Flies in Maputo, Mozambique: Effects on Enteric Pathogen Infection Risks. Environ. Sci. Technol. 57, 549–560 (2023).

49. Machdar, E., van der Steen, N. P., Raschid-Sally, L. & Lens, P. N. L. Application of Quantitative Microbial Risk Assessment to analyze the public health risk from poor drinking water quality in a low income area in Accra, Ghana. Sci. Total Environ. 449, 134–142 (2013).

50. Barnes, A. N., Davaasuren, A., Baasandagva, U. & Gray, G. C. A systematic review of zoonotic enteric parasitic diseases among nomadic and pastoral people. PLOS ONE 12, e0188809 (2017).

51. Zaba, T., Buene, D., Famba, E. & Joyeux, M. Factors associated with acute malnutrition among children 6-59 months in rural Mozambique. Matern. Child. Nutr. 17, e13060 (2020).

52. Ferdous, F. et al. Severity of Diarrhea and Malnutrition among Under Five-Year-Old Children in Rural Bangladesh. Am. J. Trop. Med. Hyg. 89, 223–228 (2013).

53. Yazew, T. & Daba, A. Associated Factors of Wasting among Infants and Young Children (IYC) in Kuyu District, Northern Oromia, Ethiopia. BioMed Res. Int. 2022, 9170322 (2022).

54. USAID Nawiri. Examining the complex dynamics influencing acute malnutrition in Turkana County – A longitudinal mixed-methods study to support community-driven activity design: Final technical report. (2024).

55. USAID Nawiri. Examining the complex dynamics influencing acute malnutrition in Samburu County – A longitudinal mixed-methods study to support community-driven activity design: Final technical report. (2024).

56. Tate, J. E. et al. Global, Regional, and National Estimates of Rotavirus Mortality in Children <5 Years of Age, 2000–2013. Clin. Infect. Dis. 62, S96–S105 (2016).

57. Kotloff, K. L. et al. Burden and aetiology of diarrhoeal disease in infants and young children in developing countries (the Global Enteric Multicenter Study, GEMS): a prospective, case-control study. The Lancet 382, 209–222 (2013).

58. Cantrell, M. E. et al. Hands Are Frequently Contaminated with Fecal Bacteria and Enteric Pathogens Globally: A Systematic Review and Meta-analysis. ACS Environ. Au 3, 123–134 (2023).

59. Pickering, A. J. et al. Hands, Water, and Health: Fecal Contamination in Tanzanian Communities with Improved, Non-Networked Water Supplies. Environ. Sci. Technol. 44, 3267–3272 (2010)

