## Supplemental Information for "Zoonotic and environmental sources of infant enteric pathogen infections identified with longitudinal sampling"

**Methods**

All samples were stored in a cooler on ice until transport to the field laboratory.

*Household survey*

In each household visited, we conducted a household survey with the child’s caregiver over 16 years of age to collect information on child health, demographics, water, sanitation, and hygiene (WASH) status, and child behavior. The child health portion of the survey was repeated at each longitudinal visit in the longitudinal cohort.

*Sample Collection*

Stool sample kits consisted of a 50ml sterile container, gloves, aluminum foil, and a spatula. Animal fecal samples were collected with sterile scoops into sterile 50 ml containers.

For soil sample collection, field workers marked a 30 cm by 30 cm area using a disposable sterile stencil and scraped the top layer of soil within the stencil into a sterile Whirlpak bag using a sterile disposable plastic scoop; the sample area was scraped once vertically and once horizontally to collect approximately 50 g of soil. Field staff prioritized sampling locations that were shaded.

Food was scooped to fill a 50mL sterile plastic tube using a sterile spoon attached to the lid of the tube.

For hand rinses, the hand was massaged from the outside of the bag for 15 seconds, followed by 15 seconds of shaking. The same procedure was repeated with the right hand in the same bag, and the rinse water was preserved in the Whirlpak bag

*Dry Weight Estimations*

The dry weight and percent moisture in soil and food samples were determined by weighing out 5g of each sample and drying in a drying oven at 110°C for 24 hours. Weights of dried samples were recorded and used to estimate the percent moisture in the samples.

*IDEXX E. coli culture*

*Culturing E. coli from environmental and fecal samples*

Stored water, source water, hand rinse, and fomite rinse samples were processed by decanting 100 mL of sample directly into IDEXX trays. Food and soil samples required a homogenization step before processing with IDEXX. Four grams of each sample were placed into a 50 mL falcon tube with 40 mL distilled water, shaken by hand for 30 seconds, and vortexed for 2 minutes. One mL of this solution was mixed with 99 mL sterile water and the resulting 100 mL were processed with IDEXX. Samples were incubated at 35°C for 24 hours to quantitatively enumerate *E. coli* and total coliforms. Field and lab blanks were processed each day, and 5% of samples were processed in duplicate for quality assurance.

All samples were preserved on ice in cooler boxes and transported to the field lab to be processed within 6 hours of sample collection using the IDEXX most probable number (MPN) method with Colilert media and QuantiTray 2000s to detect *E. coli* and total coliform. The IDEXX Quanti-Tray 2000s used for *E. coli* enumeration in this study contains 49 large wells and 48 small wells (10% the volume of large wells) and can quantify in the range of 1-2,419 colonies; input is 100 mL. After incubation for 24 hours, each well individually fluoresces if *E. coli* is present. Quantities of *E. coli* are calculated using a most probable number (MPN) approach based on the number of large and small wells that fluoresce.

The range of quantification for drinking water, source water, fomite, and hand rinse samples is 1 - 2,419 colonies. For soil and food, the range of quantification is 40 - 96,760 colonies because of soil dilution.

Field and lab blanks were processed each day, and 5% of samples were processed in duplicate for quality assurance.

Field blanks consisted of 100 mL of sterile water poured into WhirlPak bags that were transported to and from the field along with other samples. Following drinking water sample collection, enumerators were instructed to open and close the field blanks in the same manner as the main samples. Any *E. coli* detected in field blanks would indicate cross-contamination during sample collection and transport.

Laboratory blanks consisted of 100 mL sterile water processed alongside field samples for IDEXX. The sterile water was poured directly into the IDEXX trays after all other samples were processed. *E. coli* detection in laboratory blanks would indicate cross-contamination within the laboratory.

*Aliquoting for molecular analysis*

Fecal, food, and soil samples were vortexed to evenly distribute samples in the preservative. Aliquots were stored in 2mL tubes at -20°C in the local field lab before being shipped to Kenya Medical Research Institute (KEMRI) on dry ice for storage at -80°C, then shipped on dry ice to UC Berkeley for enteric pathogen molecular analysis. Water and hand rinse filters were stored in 5mL tubes with Zymo RNA/DNA shield at -20°C until transfer to KEMRI, where they were stored at -80°C.

*QA/QC*

Detection of total *E. coli* by the *uidA* assay on the TaqMan Array Card was used as an endogenous process control to confirm extraction success for fecal and soil samples. For food, drinking water, and hand rinse samples, Fisher’s xenobiotic control was spiked into samples during lysis and measured on the TaqMan Array Card to confirm extraction success, as *E. coli* was not expected to be present in all of these sample types.

*Reverse transcription*

Reverse transcription was conducted using the SuperScript IV VILO kit. The reaction consisted of 4 µL of SuperScript mastermix, 7 µL of DNA/RNA free water, and 9 µL of template (sample). Reverse transcription (RT) was performed on 9 µL of nucleic acids extract using the Invitrogen SuperScript IV VILO kit with the following cycling parameters: 25°C for 10 min, 50°C for 10 min, and 85°C for 2 min. The RT output was diluted to a total volume of 55 µL and mixed 1:1 with 55 µL of Applied Biosystems TaqMan Fast Advanced Master Mix. 100 µL of this solution was loaded onto the TAC and ran with the following cycling parameters: 50°C for 2 min, 92°C for 10 minutes, and 50 cycles of 95°C for 1 second and 60°C for 20 seconds.

Samples were then brought to 4°C before further processing.

*Preamplification*

Food, child hand rinse, and drinking water samples were subjected to preamplification prior to analysis to increase the ability to detect nucleic acids. For preamplification, we used a custom preamplification pool using the primers in our TACs (Life Sciences, USA). Reactions consisted of 20 µL of template, 10 µL of preamp pool, and 10 µL of the TaqMan Fast Advanced mastermix. Cycling parameters were 95°C for 2 min, 14 cycles of 95°C for 15 s and 60°C for 2 min, and 99°C for 10 min. Following preamplification, 10 µL of the total reaction were used as input into the TAC reaction mixture.

*TaqMan Array Card standard curve*

We used three custom gBlock DNA sequences (IDT, Iowa, USA) as template for our standard curve. Each gBlock was resuspended and all three mixed at equal concentration to create one master standard curve. We ran a 7-point standard curve on one TaqMan Array Card (TAC) with concentrations ranging from 10 gc/rxn to 10^7^gc/rxn. We repeated the standard curve across a total of 6 TACs distributed throughout the sample processing period. A master standard curve was fitted to the results for each target using a linear mix//ed effects model with batch effects for card (**Table S2**). Efficiencies ranged from 96 – 118% with a median of 108%. Standard curves were used to calculate pathogen concentrations in all sample types. For food, hand rinse, and water samples, a 10^14^ reduction in estimated quantity was applied to account for the 14 cycles of preamplification. To estimate quantities for pathogens with multiple targets, the quantity of the target with the greatest value was used.

*Limit of detection calculations*

The limit of detection (LOD) for each assay was defined as the lowest point on the standard curve where >50% of replicates amplified. The LOD for each sample type was calculated from each assay LOD using the dilution factor throughout the extraction, RT, and TAC process.

*Inhibition Testing*

Inhibition testing was conducted on soil and fecal samples, as these sample types would be the most likely to be inhibited. C_q_ values were compared between the raw and diluted (1:10, 1:20, and 1:50) sample. Samples were considered to be inhibited if the difference between the theoretical and actual C_q_ difference exceeded 1 cycle. The theoretical C_q_ difference for a raw sample *versus* a dilution is 3.32 cycles (1:10 dilution), 4.32 cycles (1:20 dilution), and 5.6 cycles (1:50 dilution).

*Poverty line calculations*

We used the Kenya Poverty Probability Index (PPI) to calculate the probability of each household falling below the poverty line and the food poverty line.^1^ Questions from the PPI index were included in our household surveys, and probabilities for poverty calculated using the methods described in the PPI report.

**Results**

*QA/QC*

Samples without amplification in the *uidA* *E. coli* gene (human stool, animal feces, and food) or the Fisher endogenous RNA control (soil, drinking water, and hand rinses) were reprocessed and if still not positive for the above controls, excluded from the analysis. In total, 33 of 871 samples were excluded. All field, laboratory, and extraction blanks were negative for total *E. coli*. Inhibition testing found no evidence of sample inhibition so undiluted samples were processed in TAC (**Table S17**).

*Household Enrollment and Characteristics*

In total, we enrolled 100 households with a child under 2 years of age (**Table S13**). Approximately 25 children were enrolled in each of the age groups: 0-2 (24 households), 3-5 (22), 6-11 (30), and 12-23 months (24). We conducted structured observations in 56 households and enrolled 60 households in the longitudinal cohort. Household surveys conducted as part of this study agree with prior surveys. We find that 64% of households fall below the Kenya poverty line and 66% below the Kenya food poverty line, indicating probable food insecurity.^1^ Eighty-six percent of households reported owning livestock, most commonly goats (83%), chickens (57%), dogs (49%), and cattle (42%). Animal feces were observed in every household, primarily sheep or goat (n=60 of 88 observations), poultry (33), cow or buffalo (17), and dog or cat (14). Ninety-three percent of households lacked electricity access and 71% of child caregivers did not complete primary school.

*WASH Access*

Access to water, sanitation, and hygiene (WASH) was limited in the study regions. Fifty-nine percent of households lacked access to any type of latrine, and only 4% reported having a handwashing station available. Latrine types used were covered pit latrines with slab (15) and without slab (9), and uncovered pit latrines with slab (4) and without slab (10); 2 households reported using a ventilated improved pit (VIP). The primary drinking water sources were piped water (n=50) and boreholes (36), with the remaining households relying on springs (5), dug wells (5), surface water (2), and vendor water (2). Sixty-six households reported that their drinking water source was not available when needed at least once in the last two weeks; 50% of these cases were due to the source running out of water, 8% due to lack of electricity or fuel, and the remaining cases for various reasons. Reported one-way walk times to the primary water source ranged from 0 to 300 minutes, with a median time of 10 minutes and a standard deviation of 46 minutes. Notably, no respondents reported treating drinking water. When households had stored water on premises at the time of our visit (n=98), the median water storage time was 6 hours.

*Child Health*

Overall, caregivers reported diarrhea in the last 7 days for over one-third of children (**Table S14**). Diarrheal prevalence peaked in the 3-5 and 6-11 month age groups. Further, diarrheal prevalence was greater in Samburu than Turkana. In our surveys we asked about diarrhea using two different definitions: 1) caregiver-defined diarrhea, 2) watery/soft stool and 3+ bowel movements in 24 hours. Diarrheal prevalence was similar between the two definitions. The overall prevalence of diarrhea in the last 7 days using each definition were 1) 36% and 2) 28%.

*Food hygiene*

For all 28 food samples collected, field staff recorded information on food storage and serving practices. Overall, 57% of food was cooled without a lid, and 64% was not reheated before serving. Food was frequently stored uncovered (46%) and on the ground (43%). Field staff observed the food preparation areas and found flies (54%), trash (43%), and animal feces (21%).

**Discussion**

We found a positive association between maternal prenatal antibiotic use and child infection. Prenatal antibiotic use can alter the maternal microbiome and, in turn, impact child microbiome and immune development after birth. Gut colonization begins in the womb, and disruptions of the maternal microbiome during pregnancy can alter the colonization period in the fetus.^2^ Prior research has shown that mother’s antibiotic use before or during pregnancy is associated with infection-related hospitalizations in children.^3^ Reducing environmental exposure to pathogens and subsequent morbidity during pregnancy could reduce unnecessary antibiotic use. Promoting antibiotic stewardship will also be vital in reducing unnecessary antibiotic use.^2^

### Tables

| Table S1. Detection of pathogens by sample type | | | | | | | | | | | | | |
| --- | --- | --- | --- | --- | --- | --- | --- | --- | --- | --- | --- | --- | --- |
|  |  | Camel Feces | Cattle Feces | Poultry Feces | Food | Child Hands | Child Stool | Dog Feces | Drinking Water | Goat Feces | Soil | Mother Stool | Sheep Feces |
| Bacteria | Aeromonas | 0 / 3 | 1 / 45 | 0 / 55 | 0 / 40 | 6 / 92 | 0 / 162 | 0 / 13 | 5 / 116 | 0 / 57 | 1 / 163 | 0 / 60 | 0 / 65 |
|  | Bacteroides fragilis | 1 / 3 | 2 / 45 | 7 / 55 | 0 / 40 | 10 / 92 | 19 / 162 | 6 / 13 | 2 / 116 | 4 / 57 | 18 / 163 | 6 / 60 | 9 / 65 |
|  | Campylobacter coli/jejuni | 0 / 3 | 1 / 45 | 14 / 55 | 0 / 40 | 1 / 92 | 16 / 162 | 0 / 13 | 0 / 116 | 4 / 57 | 17 / 163 | 3 / 60 | 3 / 65 |
|  | Clostridium difficile | 0 / 3 | 0 / 45 | 0 / 55 | 0 / 40 | 0 / 92 | 2 / 162 | 1 / 13 | 0 / 116 | 0 / 57 | 0 / 163 | 0 / 60 | 0 / 65 |
|  | E. coli/ Shigella | 1 / 3 | 0 / 45 | 3 / 55 | 1 / 40 | 11 / 92 | 3 / 162 | 3 / 13 | 1 / 116 | 0 / 57 | 11 / 163 | 13 / 60 | 0 / 65 |
|  | EAEC | 0 / 3 | 0 / 45 | 26 / 55 | 0 / 40 | 13 / 92 | 91 / 162 | 7 / 13 | 0 / 116 | 8 / 57 | 68 / 163 | 37 / 60 | 10 / 65 |
|  | EHEC 0157H7 | 0 / 3 | 0 / 45 | 3 / 55 | 0 / 40 | 2 / 92 | 3 / 162 | 0 / 13 | 0 / 116 | 0 / 57 | 4 / 163 | 2 / 60 | 1 / 65 |
|  | EIEC | 1 / 3 | 0 / 45 | 3 / 55 | 0 / 40 | 1 / 92 | 0 / 162 | 2 / 13 | 0 / 116 | 0 / 57 | 3 / 163 | 11 / 60 | 0 / 65 |
|  | EPEC | 0 / 3 | 2 / 45 | 32 / 55 | 2 / 40 | 22 / 92 | 59 / 162 | 9 / 13 | 8 / 116 | 13 / 57 | 61 / 163 | 30 / 60 | 15 / 65 |
|  | ETEC-LT | 0 / 3 | 1 / 45 | 18 / 55 | 2 / 40 | 42 / 92 | 24 / 162 | 4 / 13 | 14 / 116 | 6 / 57 | 59 / 163 | 17 / 60 | 6 / 65 |
|  | ETEC-ST | 0 / 3 | 0 / 45 | 7 / 55 | 0 / 40 | 25 / 92 | 3 / 162 | 2 / 13 | 4 / 116 | 2 / 57 | 25 / 163 | 3 / 60 | 1 / 65 |
|  | Helicobacter pylori | 0 / 3 | 0 / 45 | 0 / 55 | 0 / 40 | 2 / 92 | 10 / 162 | 0 / 13 | 0 / 116 | 0 / 57 | 0 / 163 | 6 / 60 | 0 / 65 |
|  | STEC | 1 / 3 | 10 / 45 | 19 / 55 | 1 / 40 | 17 / 92 | 12 / 162 | 6 / 13 | 7 / 116 | 29 / 57 | 19 / 163 | 8 / 60 | 34 / 65 |
|  | Salmonella enterica | 0 / 3 | 0 / 45 | 2 / 55 | 0 / 40 | 2 / 92 | 1 / 162 | 0 / 13 | 2 / 116 | 0 / 57 | 7 / 163 | 0 / 60 | 0 / 65 |
|  | Salmonella typhi | 0 / 3 | 0 / 45 | 0 / 55 | 0 / 40 | 0 / 92 | 0 / 162 | 0 / 13 | 0 / 116 | 0 / 57 | 3 / 163 | 0 / 60 | 0 / 65 |
|  | Shigella flexneri 6 | 0 / 3 | 0 / 45 | 0 / 55 | 0 / 40 | 0 / 92 | 1 / 162 | 0 / 13 | 0 / 116 | 0 / 57 | 0 / 163 | 2 / 60 | 0 / 65 |
|  | Vibrio cholerae | 0 / 3 | 0 / 45 | 0 / 55 | 1 / 40 | 5 / 92 | 0 / 162 | 0 / 13 | 1 / 116 | 0 / 57 | 27 / 163 | 1 / 60 | 0 / 65 |
| Helminths | Ancyclostoma duodenale | 0 / 3 | 0 / 45 | 0 / 55 | 0 / 40 | 0 / 92 | 0 / 162 | 0 / 13 | 0 / 116 | 0 / 57 | 0 / 163 | 0 / 60 | 0 / 65 |
|  | Ascaris | 0 / 3 | 0 / 45 | 0 / 55 | 0 / 40 | 0 / 92 | 0 / 162 | 0 / 13 | 0 / 116 | 0 / 57 | 0 / 163 | 0 / 60 | 0 / 65 |
|  | Necator americanus | 0 / 3 | 0 / 45 | 0 / 55 | 0 / 40 | 0 / 92 | 0 / 162 | 0 / 13 | 0 / 116 | 0 / 57 | 0 / 163 | 0 / 60 | 0 / 65 |
|  | Stongyloides stercoralis | 0 / 3 | 0 / 45 | 0 / 55 | 0 / 40 | 0 / 92 | 0 / 162 | 0 / 13 | 0 / 116 | 0 / 57 | 0 / 163 | 0 / 60 | 0 / 65 |
|  | Trichuris | 1 / 3 | 1 / 45 | 0 / 55 | 0 / 40 | 0 / 92 | 0 / 162 | 0 / 13 | 0 / 116 | 7 / 57 | 5 / 163 | 0 / 60 | 7 / 65 |
| Protozoa | C hominis | 0 / 3 | 0 / 45 | 0 / 55 | 0 / 40 | 0 / 92 | 3 / 162 | 0 / 13 | 0 / 116 | 0 / 57 | 0 / 163 | 0 / 60 | 0 / 65 |
|  | C parvum | 0 / 3 | 0 / 45 | 0 / 55 | 0 / 40 | 0 / 92 | 0 / 162 | 0 / 13 | 0 / 116 | 0 / 57 | 0 / 163 | 0 / 60 | 0 / 65 |
|  | Cryptosporidium spp. | 0 / 3 | 17 / 45 | 13 / 55 | 0 / 40 | 8 / 92 | 6 / 162 | 5 / 13 | 14 / 116 | 10 / 57 | 57 / 163 | 1 / 60 | 10 / 65 |
|  | Cyclospora cayetanesis | 0 / 3 | 0 / 45 | 0 / 55 | 0 / 40 | 2 / 92 | 0 / 162 | 1 / 13 | 0 / 116 | 2 / 57 | 5 / 163 | 0 / 60 | 3 / 65 |
|  | E. histolytica | 0 / 3 | 0 / 45 | 0 / 55 | 0 / 40 | 0 / 92 | 0 / 162 | 0 / 13 | 0 / 116 | 0 / 57 | 0 / 163 | 0 / 60 | 0 / 65 |
|  | Giardia duodenalis | 0 / 3 | 5 / 45 | 15 / 55 | 18 / 40 | 68 / 92 | 32 / 162 | 9 / 13 | 53 / 116 | 11 / 57 | 69 / 163 | 16 / 60 | 12 / 65 |
| Viruses | Adenovirus 40/41 | 0 / 3 | 0 / 45 | 3 / 55 | 0 / 40 | 14 / 92 | 24 / 162 | 4 / 13 | 6 / 116 | 0 / 57 | 8 / 163 | 5 / 60 | 2 / 65 |
|  | Astrovirus | 0 / 3 | 0 / 45 | 2 / 55 | 0 / 40 | 3 / 92 | 15 / 162 | 0 / 13 | 0 / 116 | 0 / 57 | 4 / 163 | 0 / 60 | 0 / 65 |
|  | Enterovirus | 0 / 3 | 0 / 45 | 15 / 55 | 0 / 40 | 11 / 92 | 76 / 162 | 3 / 13 | 0 / 116 | 2 / 57 | 25 / 163 | 14 / 60 | 3 / 65 |
|  | Norovirus GI | 0 / 3 | 0 / 45 | 0 / 55 | 0 / 40 | 0 / 92 | 8 / 162 | 0 / 13 | 0 / 116 | 0 / 57 | 1 / 163 | 1 / 60 | 0 / 65 |
|  | Norovirus GII | 0 / 3 | 0 / 45 | 4 / 55 | 0 / 40 | 3 / 92 | 20 / 162 | 5 / 13 | 0 / 116 | 0 / 57 | 3 / 163 | 4 / 60 | 0 / 65 |
|  | Rotavirus A | 0 / 3 | 0 / 45 | 1 / 55 | 0 / 40 | 5 / 92 | 4 / 162 | 0 / 13 | 1 / 116 | 0 / 57 | 3 / 163 | 0 / 60 | 0 / 65 |

| **Table S2.** Unadjusted risk factor analysis for pathogenic *E. coli* carriage. Presented are odds ratios of child pathogen carriage given household factors. For each risk factor, the odds ratio was calculated using logit-link models. | | | | | |
| --- | --- | --- | --- | --- | --- |
|  | Any pathogen | E. coli | Bacteria | Virus | Protozoa or Helminth |
| Probability of falling below poverty line | 3.78 (0.65, 22.06) | 4.7 (0.72, 30.86) | 4.87 (0.72, 33.03) | 1.58 (0.35, 7.09) | 0.94 (0.14, 6.43) |
| Consumes breastmilk | - | 0.5 (0.09, 2.83) | 0.23 (0.03, 2.1) | 0.58 (0.13, 2.64) | **0.15 (0.04, 0.64)** |
| Consumes food | **12.17 (1.62, 91.18)** | **4.81 (1.46, 15.82)** | **7.52 (1.69, 33.42)** | **3.28 (1.33, 8.08)** | **15.15 (5.13, 44.77)** |
| Crawls | **2.3x10^7^ (1.1x10^7^, 4.8x10^7^)** | 2.1 (0.63, 6.97) | 2.65 (0.71, 9.97) | 2.55 (0.86, 7.53) | **10.67 (3.16, 35.95)** |
| Walks | **7.4x10^6^ (3.3x10^6^, 1.6x10^7^)** | 1.46 (0.35, 5.98) | 5.24 (0.63, 43.81) | **8.41 (1.02, 69.5)** | **8.85 (2.59, 30.2)** |
| Mother consumed antibiotics during pregnancy | 1.83 (0.59, 5.71) | **2.54 (1.03, 6.28)** | **3.0 (1.17, 7.7)** | 0.68 (0.31, 1.49) | 1.4 (0.54, 3.68) |
| Walk time to water source | 1.02 (0.98, 1.07) | 1.01 (0.97, 1.06) | **1.05 (1.0, 1.1)** | 1.01 (0.99, 1.03) | **1.03 (1.0, 1.05)** |
| Latrine access | 0.54 (0.19, 1.57) | 0.79 (0.32, 1.97) | 0.8 (0.32, 2.02) | 0.77 (0.35, 1.67) | 1.33 (0.5, 3.5) |
| Owns livestock | 1.34 (0.36, 5.05) | 1.76 (0.6, 5.19) | 1.9 (0.64, 5.66) | 0.91 (0.28, 2.9) | 2.37 (0.6, 9.43) |
| Owns cattle | 0.3 (0.09, 1.05) | 0.5 (0.18, 1.41) | 0.42 (0.14, 1.23) | 0.81 (0.33, 1.96) | 0.62 (0.21, 1.8) |
| Owns poultry | 0.92 (0.32, 2.67) | 0.92 (0.37, 2.31) | 1.04 (0.41, 2.63) | 1.33 (0.58, 3.07) | 1.21 (0.46, 3.18) |
| Owns dogs | 0.47 (0.14, 1.65) | 0.81 (0.28, 2.29) | 0.68 (0.23, 2.03) | 0.59 (0.25, 1.42) | 0.72 (0.25, 2.06) |
| Insects in food preparation area | 0.53 (0.19, 1.51) | 0.67 (0.27, 1.66) | 0.6 (0.23, 1.52) | **0.44 (0.2, 0.99)** | 0.65 (0.24, 1.74) |
| Dispose of animal feces in compound | 1.34 (0.44, 4.02) | 2.25 (0.82, 6.16) | 2.33 (0.82, 6.63) | 0.98 (0.41, 2.31) | 0.61 (0.22, 1.74) |

| **Table S3.** Adjusted risk factor analysis for pathogenic *E. coli* carriage. Presented are odds ratios of child pathogen carriage given household factors. For each risk factor, an adjusted odds ratio was calculated using logit-link models, controlling for child age category. | | | |
| --- | --- | --- | --- |
| Risk Factor | EAEC | EPEC | ETEC-LT |
| Probability of falling below poverty line | 3.04 (0.61, 15.16) | 5.85 (0.94, 36.19) | 5.09 (0.88, 29.42) |
| Consumes breastmilk | 1.21 (0.22, 6.59) | 0.19 (0.03, 1.04) | 1.86 (0.24, 14.5) |
| Consumes food | **3.37 (1.07, 10.66)** | 1.34 (0.56, 3.23) | 3.81 (0.76, 19.17) |
| Crawls | 0.22 (0.04, 1.2) | 0.97 (0.27, 3.49) | 2.32 (0.49, 10.84) |
| Walks | **0.12 (0.04, 0.34)** | **4.4x10^6^ (9.0x10^5^, 2.1x10^7^)** | **8.6 (3.01, 24.54)** |
| Mother consumed antibiotics during pregnancy | **2.68 (1.22, 5.89)** | 0.88 (0.39, 1.97) | **3.81 (1.13, 12.85)** |
| Walk time to water source | 1.0 (0.98, 1.02) | 1.04 (0.94, 1.16) | 1.0 (0.98, 1.02) |
| Latrine access | 0.88 (0.39, 2.02) | **0.42 (0.2, 0.88)** | 0.91 (0.30, 2.73) |
| Owns livestock | 0.9 (0.32, 2.52) | 1.84 (0.61, 5.53) | 2.17 (0.40, 11.65) |
| Owns cattle | 0.4 (0.16, 1.01) | 0.70 (0.27, 1.83) | 0.88 (0.26, 2.96) |
| Owns poultry | 0.57 (0.25, 1.28) | 1.35 (0.57, 3.2) | 2.04 (0.66, 6.25) |
| Owns dogs | 0.65 (0.25, 1.67) | 1.26 (0.53, 2.99) | 0.44 (0.15, 1.33) |
| Insects in food preparation area | 0.55 (0.24, 1.26) | 1.58 (0.67, 3.71) | 0.79 (0.27, 2.32) |
| Disposes of animal feces in compound | 1.23 (0.54, 2.84) | 2.23 (0.97, 5.14) | 2.0 (0.66, 6.06) |

| **Table S4.** Unadjusted risk factor analysis for pathogenic *E. coli* carriage. Presented are odds ratios of child pathogen carriage given household factors. For each risk factor, the odds ratio was calculated using logit-link models. | | | |
| --- | --- | --- | --- |
| Risk Factor | EAEC | EPEC | ETEC-LT |
| Age group |  |  |  |
| 0-2 months | Ref. | Ref. | Ref. |
| 3-5 months | 1.1 (0.42, 2.87) | 1.87 (0.7, 4.99) | **6.83 (1.38, 33.87)** |
| 6-11 months | **6.51 (2.15, 19.65)** | **4.71 (1.58, 14.1)** | 4.94 (0.95, 25.7) |
| 12-23 months | 1.15 (0.32, 4.12) | 2.24 (0.54, 9.26) | **28 (4.71, 166.29)** |
| Probability of falling below poverty line | 1.54 (0.31, 7.58) | 2.76 (0.54, 14.04) | 3.19 (0.53, 19.26) |
| Consumes breastmilk | 0.75 (0.19, 2.98) | **0.21 (0.06, 0.74)** | 0.43 (0.10, 1.87) |
| Consumes food | **3.65 (1.53, 8.73)** | **2.28 (1.0, 5.21)** | **5.61 (1.92, 16.37)** |
| Crawls | 1.36 (0.56, 3.26) | 1.97 (0.88, 4.39) | **3.93 (1.3, 11.86)** |
| Walks | 0.62 (0.21, 1.86) | 1.97 (0.57, 6.87) | **8.99 (2.75, 29.43)** |
| Mother consumed antibiotics during pregnancy | **2.84 (1.28, 6.31)** | 1.01 (0.46, 2.19) | **2.95 (1.08, 8.08)** |
| Walk time to water source | 1.0 (0.98, 1.01) | 1.03 (0.98, 1.08) | 1.0 (0.98, 1.02) |
| Latrine access | 1.08 (0.47, 2.46) | 0.56 (0.26, 1.18) | 0.99 (0.36, 2.74) |
| Owns livestock | 1.21 (0.43, 3.37) | 2.38 (0.75, 7.51) | 2.83 (0.61, 13.01) |
| Owns cattle | 0.42 (0.17, 1.04) | 0.67 (0.26, 1.69) | 0.63 (0.21, 1.85) |
| Owns poultry | 0.77 (0.34, 1.75) | 1.57 (0.70, 3.51) | 1.97 (0.69, 5.59) |
| Owns dogs | 0.52 (0.21, 1.3) | 1.02 (0.42, 2.45) | 0.33 (0.11, 1.01) |
| Insects in food preparation area | 0.61 (0.27, 1.36) | 1.32 (0.57, 3.06) | 0.54 (0.18, 1.62) |
| Disposes of animal feces in compound | 1.27 (0.54, 2.97) | 2.23 (0.99, 5.04) | 1.86 (0.64, 5.4) |

| **Table S5.** Unadjusted risk factor analysis for adenovirus, enterovirus, giardia, and norovirus GII. Presented are odds ratios of child pathogen carriage given household factors. For each risk factor, an unadjusted odds ratio was calculated using logit-link models. | | | | |
| --- | --- | --- | --- | --- |
| Risk Factor | Adenovirus | Enterovirus | Giardia duodenalis | Norovirus GII |
| Age group |  |  |  |  |
| 0-2 months | Ref. | Ref. | Ref. | Ref. |
| 3-5 months | **8.6x10^7^ (3.5x10^7^, 2.1x10^8^)** | 1.98 (0.77, 5.04) | **17.54 (2.05, 150.18)** | **10.6 (1.33, 84.33)** |
| 6-11 months | **7.9x10^7^ (3.1x10^7^, 2.0x10^8^)** | 2.22 (0.83, 5.96) | **21.62 (2.48, 188.46)** | **14.25 (1.63, 124.79)** |
| 12-23 months | **2.2x10^8^ (6.9x10^7^, 7.3x10^8^)** | **24.44 (2.89, 207.04)** | **171 (16.67, 1754.41)** | **19 (1.85, 194.93)** |
| Probability of falling below poverty line | 0.53 (0.08, 3.59) | 1.05 (0.26, 4.29) | 0.51 (0.07, 3.69) | 8.38 (0.37, 190.76) |
| Consumes breastmilk | 1.81 (0.20, 16.69) | 0.45 (0.07, 2.77) | **0.18 (0.04, 0.95)** | 0.61 (0.12, 3.08) |
| Consumes food | **5.61 (1.71, 18.42)** | **3.39 (1.42, 8.08)** | **14.57 (4.35, 48.83)** | 2.36 (0.76, 7.31) |
| Crawls | 3.31 (0.95, 11.57) | 2.41 (0.78, 7.46) | **12.43 (3.41, 45.38)** | 1.79 (0.56, 5.75) |
| Walks | 2.84 (0.72, 11.25) | **17.11 (2.08, 141.06)** | **7.87 (1.93, 32.07)** | **3.67 (1.03, 13.08)** |
| Mother consumed antibiotics during pregnancy | 0.77 (0.25, 2.4) | 1.19 (0.55, 2.59) | 2.05 (0.71, 5.96) | 0.66 (0.22, 1.96) |
| Walk time to water source | 1.01 (0.99, 1.03) | 0.99 (0.97, 1.02) | 1.02 (1.0, 1.04) | **1.02 (1.00, 1.03)** |
| Latrine access | 1.21 (0.41, 3.61) | 0.95 (0.44, 2.06) | 1.54 (0.54, 4.45) | 1.91 (0.66, 5.52) |
| Owns livestock | 1.72 (0.31, 9.49) | 0.94 (0.35, 2.48) | 1.85 (0.45, 7.52) | 4.92 (0.66, 36.9) |
| Owns cattle | 0.69 (0.21, 2.22) | **0.27 (0.11, 0.66)** | 0.55 (0.17, 1.81) | 2.29 (0.73, 7.19) |
| Owns poultry | 1.3 (0.43, 3.96) | 1.28 (0.58, 2.82) | 1.49 (0.52, 4.32) | 1.78 (0.58, 5.48) |
| Owns dogs | 0.46 (0.14, 1.55) | 0.7 (0.29, 1.69) | 0.81 (0.25, 2.59) | 1.98 (0.61, 6.4) |
| Insects in food preparation area | 0.68 (0.21, 2.18) | 0.43 (0.19, 0.95) | 0.56 (0.19, 1.67) | 0.95 (0.29, 3.17) |
| Disposes of animal feces in compound | 0.88 (0.27, 2.93) | 1.4 (0.6, 3.26) | 0.53 (0.18, 1.6) | 3.55 (0.97, 12.97) |

| **Table S6.** Adjusted risk factor analysis for adenovirus, enterovirus, giardia, and norovirus GII. Presented are odds ratios of child pathogen carriage given household factors. For each risk factor, an adjusted odds ratio was calculated using logit-link models and controlling for child age category. | | | | |
| --- | --- | --- | --- | --- |
| Risk Factor | Adenovirus | Enterovirus | Giardia duodenalis | Norovirus GII |
| Probability of falling below poverty line | 0.96 (0.13, 6.87) | 1.45 (0.31, 6.79) | 0.87 (0.11, 7.13) | 17.58 (0.80, 388.21) |
| Consumes breastmilk | **12.57 (1.1, 144.13)** | 2.19 (0.16, 29.56) | 0.92 (0.05, 16.04) | 1.52 (0.22, 10.44) |
| Consumes food | 2.84 (0.61, 13.16) | 2.24 (0.78, 6.47) | **8.15 (1.84, 36.06)** | 0.99 (0.30, 3.28) |
| Crawls | 1.79 (0.35, 9.17) | 0.93 (0.18, 4.84) | **9.57 (1.76, 52.05)** | 0.57 (0.12, 2.63) |
| Walks | - | 6.18 (0.70, 54.26) | - | **5.12 (1.77, 14.8)** |
| Mother consumed antibiotics during pregnancy | 0.73 (0.16, 3.29) | 1.11 (0.47, 2.63) | 2.6 (0.65, 10.32) | 0.55 (0.16, 1.88) |
| Walk time to water source | 1.01 (0.99, 1.03) | 0.99 (0.97, 1.01) | 1.02 (0.99, 1.05) | **1.01 (1.00, 1.03)** |
| Latrine access | 1.03 (0.31, 3.39) | 0.87 (0.37, 2.03) | 1.46 (0.44, 4.91) | 1.65 (0.59, 4.65) |
| Owns livestock | 1.09 (0.17, 6.87) | 0.7 (0.26, 1.89) | 1.09 (0.21, 5.61) | 3.49 (0.50, 24.42) |
| Owns cattle | 0.91 (0.25, 3.26) | **0.32 (0.13, 0.80)** | 0.85 (0.20, 3.64) | **3.42 (1.09, 10.74)** |
| Owns poultry | 1.19 (0.37, 3.84) | 1.2 (0.53, 2.73) | 1.35 (0.41, 4.42) | 1.59 (0.54, 4.67) |
| Owns dogs | 0.72 (0.21, 2.48) | 0.96 (0.38, 2.39) | 1.46 (0.36, 5.95) | **3.3 (1.01, 10.8)** |
| Insects in food preparation area | 1.09 (0.34, 3.49) | 0.49 (0.21, 1.13) | 0.84 (0.25, 2.78) | 1.35 (0.42, 4.36) |
| Disposes of animal feces in compound | 0.81 (0.24, 2.76) | 1.44 (0.60, 3.46) | 0.44 (0.12, 1.62) | 3.61 (1.04, 12.46) |

| **Table S7**. Correlations between pathogens and total E. coli measured by TAC (*uidA* gene): *r* correlation, p value. | | | | | | | | |
| --- | --- | --- | --- | --- | --- | --- | --- | --- |
| **Pathogen** | **Soil** | **Food** | **Hands** | **Drinking water** | **Chicken feces** | **Goat feces** | **Sheep feces** | **Child stool** |
| Adenovirus 40/41 | 0.05, 0.94 |  |  |  |  | 0.23, 0.94 | -0.16, 0.94 |  |
| Astrovirus | 0.39, 0.66 |  |  |  | -0.1, 0.94 |  |  | -0.18, 0.94 |
| Clostridium difficile |  |  |  |  |  |  |  | -0.09, 0.94 |
| Cryptosporidium spp. | -0.08, 0.94 |  | 0.07, 0.94 |  | -0.08, 0.94 |  |  | -0.01, 0.94 |
| Cyclospora cayetanesis | -0.07, 0.94 |  |  |  | **0.78, 0.00** |  |  | 0.26, 0.37 |
| E. coli/ Shigella | 0.06, 0.94 |  | 0.45, 0.21 |  |  |  |  |  |
| E. histolytica | -0.17, 0.94 |  |  |  |  |  |  | 0.49, 0.21 |
| Enterovirus | 0.36, 0.94 |  |  |  |  |  |  |  |
| Giardia duodenalis | 0.08, 0.94 |  | **0.68, 0.00** | **1, 0.00** | 0.22, 0.94 |  |  | -0.1, 0.94 |
| Rotavirus A | -0.07, 0.94 |  | -0.18, 0.94 |  | -0.1, 0.94 | **0.71, 0.00** | -0.05, 0.94 | 0.45, 0.21 |
| Salmonella typhi | 0.01, 0.94 | -0.18, 0.94 | 0.01, 0.94 | 0.1, 0.94 | 0.02, 0.94 | 0.05, 0.94 | -0.21, 0.94 | -0.08, 0.94 |
| Stongyloides stercoralis | 0.07, 0.94 |  |  |  |  |  |  |  |

| **Table S8.** Correlations between pathogens measured by TAC and total *E. coli* measured in culture; correlations between detection (right) and quantities (left). Displayed is the Pearon’s r, p value adjusted for multiple comparisons using the Bonferroni adjustment. Bold indicates statistical significant (p < 0.05) | | | | | | | | | |
| --- | --- | --- | --- | --- | --- | --- | --- | --- | --- |
|  | **Positive/negative** | | | |  | **Quantities** | | | |
| **Pathogen** | **Soil** | **Food** | **Hands** | **Drinking water** |  | **Soil** | **Food** | **Hands** | **Drinking water** |
| Adenovirus 40/41 | 0.13, 0.72 |  | -0.04, 0.93 | 0.18, 0.72 |  | 0.17, 0.91 |  |  |  |
| Aeromonas | 0.09, 0.72 |  | 0.1, 0.80 | -0.04, 0.90 |  | -0.08, 0.91 |  | 0.25, 0.91 | -0.03, 0.92 |
| Ancyclostoma duode le |  |  |  |  |  | -0.05, 0.91 |  |  |  |
| Ascaris |  |  |  |  |  |  |  |  |  |
| Astrovirus |  |  | -0.13, 0.72 |  |  |  |  | -0.04, 0.91 |  |
| C hominis |  |  |  |  |  |  |  |  |  |
| C parvum |  |  |  |  |  |  |  |  |  |
| Campylobacter coli/jejuni | -0.02, 0.94 |  | -0.13, 0.72 |  |  | -0.04, 0.91 |  | 0.18, 0.91 | -0.06, 0.91 |
| Clostridium difficile |  |  |  |  |  |  |  |  |  |
| Cryptosporidium spp. | 0.18, 0.72 |  | 0.19, 0.72 | -0.05, 0.90 |  |  |  |  |  |
| Cyclospora cayetanesis | -0.07, 0.80 |  | 0.32, 0.72 |  |  | -0.08, 0.91 |  | 0.34, 0.32 |  |
| E. coli/ Shigella | 0.13, 0.72 |  | 0.28, 0.72 |  |  | 0.10, 0.91 |  | 0.41, 0.13 |  |
| E. histolytica |  |  |  |  |  |  |  |  |  |
| EAEC | 0.33, 0.34 |  | -0.09, 0.80 |  |  | 0.15, 0.91 |  | 0.21, 0.91 |  |
| EHEC 0157H7 | 0.06, 0.81 |  |  |  |  |  |  |  |  |
| EIEC | 0.16, 0.72 |  | 0.22, 0.72 |  |  | -0.08, 0.91 |  | **0.47, 0.05** |  |
| EPEC | 0.13, 0.72 | -0.35, 0.72 | -0.09, 0.80 | 0.09, 0.80 |  | -0.04, 0.91 |  | 0.11, 0.91 |  |
| ETEC-LT | 0.18, 0.72 | 0.29, 0.72 | -0.04, 0.93 | 0.08, 0.80 |  | 0.01, 0.96 | -0.15, 0.91 | 0.09, 0.91 | **0.56, 0.00** |
| ETEC-ST | 0.08, 0.78 |  | -0.14, 0.72 | -0.25, 0.72 |  | -0.06, 0.91 |  | -0.07, 0.91 | -0.06, 0.91 |
| Enterovirus | 0.12, 0.72 |  | -0.14, 0.72 |  |  | 0.03, 0.91 |  | -0.07, 0.91 |  |
| Giardia duodenalis | 0.13, 0.72 | -0.35, 0.72 | -0.17, 0.72 | -0.15, 0.72 |  | -0.04, 0.91 | -0.24, 0.91 | -0.08, 0.91 | -0.02, 0.92 |
| Helicobacter pylori |  |  | -0.13, 0.72 |  |  |  |  | -0.04, 0.91 |  |
| Necator americanus |  |  |  |  |  |  |  |  |  |
| Norovirus GI |  |  |  |  |  |  |  |  |  |
| Norovirus GII | 0.09, 0.72 |  | -0.02, 0.95 |  |  | **0.42, 0.01** |  | **0.47, 0.05** |  |
| Rotavirus A | 0.01, 0.95 |  | 0.32, 0.72 |  |  | 0.03, 0.91 |  | 0.02, 0.94 |  |
| STEC | -0.01, 0.96 |  | 0.00, 0.98 | 0.05, 0.90 |  | 0.05, 0.91 |  |  |  |
| Salmonella enterica |  |  | -0.13, 0.72 |  |  |  |  | -0.04, 0.91 |  |
| Salmonella typhi | -0.05, 0.86 |  |  |  |  | -0.05, 0.91 |  |  |  |
| Shigella flexneri 6 |  |  |  |  |  |  |  |  |  |
| Stongyloides stercoralis |  |  |  |  |  |  |  |  |  |
| Trichuris | -0.16, 0.72 |  |  |  |  | -0.05, 0.91 |  |  |  |
| Vibrio cholerae | 0.23, 0.72 |  | -0.13, 0.72 |  |  | -0.07, 0.91 |  | -0.04, 0.91 |  |

| **Table S9:** Primers and probes used in TaqMan Array Card | | | | | | |
| --- | --- | --- | --- | --- | --- | --- |
| **Type** | **Pathogen** | **Gene** | **Forward primer** | **Reverse primer** | **Probe** | **Source** |
| Bacteria | Aeromonas | aha1 | ACCGCTGCTCATTACTCTGATG | CCAACCCAGACGGGAAGAA | TGATGGTGAGCTGGTTG | ^6^ |
|  | Campylobacter coli/jejuni | cadF | CTGCTAAACCATAGAAATAAAATTTCTCAC | CTTTGAAGGTAATTTAGATATGGATAATCG | CATTTTGACGATTTTTGGCTTGA | ^7^ |
|  | Campylobacter jejuni | hipO | CTTGCGGTCATGATGGACATAC | AGCACCACCCAAACCCTCTTCA | TGCTTGCTGCAAAGTATT | Levy |
|  | C. coli | GlyA | AAACCAAAGCTTATCGTGTGC | AGTGCAGCAATGTGTGCAAT | TAAGCTCCAACTTCATCCG | Levy |
|  | E. coli/ Shigella | ipaH | CCTTTTCCGCGTTCCTTGA | CGGAATCCGGAGGTATTGC | CGCCTTTCCGATACCGTCTCTGCA | ^7^ |
|  | EAEC | aggR | GGAAGCAATACATATCTTAGAAATGAACTC | TCGGACAACTGCAAGCATCTAC | TCCGTATATTATCATCAGGGCATCCTTTAGGCGT | ^8^ |
|  | EAEC | aaiC | ATTGTCCTCAGGCATTTCAC | ACGACACCCCTGATAAACAA | TAGTGCATACTCATCATTTAAG | ^7^ |
|  | EAEC | aatA | CTGGCGAAAGACTGTATCAT | TTTTGCTTCATAAGCCGATAGA | TGGTTCTCATCTATTACAGACAGC | ^7^ |
|  | Bacteroides fragilis | bft | GGGACAAGGATTCTACCAGCTTTATA | ATTCGGCAATCTCATTCATCATT | CAATGGCGAATCCATCAG | ^9^ |
|  | Clostridium difficile | tcdB | GGTATTACCTAATGCTCCAAATAG | TTTGTGCCATCATTTTCTAAGC | CCTGGTGTCCATCCTGTTTC | ^10^ |
|  | EPEC | bfpA | TGGTGCTTGCGCTTGCT | CGTTGCGCTCATTACTTCTG | CAGTCTGCGTCTGATTCCAA | ^7^ |
|  | EPEC | eae | CATTGATCAGGATTTTTCTGGTGATA | CTCATGCGGAAATAGCCGTTA | ATACTGGCGAGACTATTTCAA | ^7^ |
|  | EIEC | virF | TCTGAGGAGGAGGTTTCTATCGATT | GAAACAGCTGATAAAAGGCAAGCT | CCGAAAGGCATCTCTTTT | Levy |
|  | ETEC-LT | LT | TTCCCACCGGATCACCAA | CAACCTTGTGGTGCATGATGA | CTTGGAGAGAAGAACCCT | ^7^ |
|  | ETEC-ST | estA | AAGCATGAATAGTAGCAATTACTGCT | TTAATAGCACCCGGTACAAGCA | AACAACACAATTCAC | ^11^ |
|  | EHEC 0157H7 | rfbE | TTTCACACTT ATTGGATGG TCTCAA | CGATGAGTTT ATCTGCAAG GTGAT | CTCTCTTTCCT CTGCGGTCCT | ^12^ |
|  | Salmonella enterica | ttr | CTCACCAGGAGATTACAACATGG | AGCTCAGACCAAAAGTGACCATC | CACCGACGGCGAGACCGACTTT | ^13^ |
|  | Salmonella typhi | tviB | TGTGGTAAAGGAACTCGGTAAA | GACTTCCGATACCGGGATAATG | TGGATGCCGAAGAGGTAAGACGAGA | Levy |
|  | Salmonella typhi | STY0201 | CGCGAAGTCAGAGTCGACATAG | AAGACCTCAACGCCGATCAC | CAGCCTGCTCCAGAACA | Levy |
|  | Shiga-like toxin 1 | stx1 | CATCGCGAGTTGCCAGAAT | GCGTAATCCCACGGACTCTTC | CTGCCGGACACATAGAAGGAAACTCATCA | ^14^ |
|  | Shiga-like toxin 2 | stx2 | CCACATCGGTGTCTGTTATTAACC | GGTCAAAACGCGCCTGATAG | TTGCTGTGGATATACGAGG | ^7^ |
|  | Shigella flexneri 6 | T3RE | CTTTCAACGCACGAATATCAAC | GAACCTGATCCAGACGGAGA | TTCTTCAGAACCGGGTTTTG | ^9^ |
|  | Shigella flexneri 6 | O-antigen | CTCCTATCCGTGATTATAGTGCA | GCACACACAACTCACTGTATTT | TCCTTCTCACGATTAAAATC | ^9^ |
|  | Vibrio cholerae | toxR | GTTTGGCGAGAGCAAGGTTT | TCTCTTCTTCAACCGTTTCCA | CGCAGAGTCGAAATGGCTTGG | ^7^ |
|  | E. coli | uidA | CGGAAGCAACGCGTAAACTC | TGAGCGTCGCAGAACATTACA | CGCGTCCGATCACCTGCGTC | ^15^ |
|  | Helicobacter pylori | ureC | GACACCAGAAAAAGCGGCTA | AGCGCATGTCTTCGGTTAAA | TCACTAAAGCGTTTTCTACC | ^10^ |
| Viruses | Adenovirus | Hexon | GCCACGGTGGGGTTTCTAAACTT | GCCCCAGTGGTCTTACATGCACATC | TGCACCAGACCCGGGCTCAG | ^7^ |
|  | Adenovirus 40/41 | Fiber | AACTTTCTCT CTTAATAGA CGCC | AGGGGGCTA GAAAACAAA A | CTGACACGGG CACTCT | ^12^ |
|  | Astrovirus | Capsid | CAGTTGCTT GCTGCGTTC A | CTTGCTAGCC ATCACACTTC T | CACAGAAGA GCAACTCCAT CGC | ^12^ |
|  | Enterovirus | 5' UTR | CCCTGAATGCGGCTAATCC | GCGATTGTCACCATWAGCAG | CCGACTACTTTGGGWGTCCGT | Levy |
|  | Norovirus GI | ORF1-ORF2 | CGYTGGATG CGNTTYCAT GA | CTTAGACGCC ATCATCATTY AC | TGGACAGGAG ATCGC | ^12^ |
|  | Norovirus GII | ORF1-ORF2 | CARGARBCNATGTTYAGR TGGATGAG | TCGACGCCATCTTCATTCACA | TGGGAGGGCGATCGCAATCT | ^7^ |
|  | Rotavirus A | NSP3 | ACCATCTWCACRTRACCCTCTATGAG | GGTCACATAACGCCCCTATAGC | AGTTAAAAGCTAACACTGTCAAA | ^7^ |
| Protozoa | Cryptosporidium spp. | 18S | GGGTTGTATTTATTAGATAAAGAACCA | AGGCCAATACCCTACCGTCT | TGACATATCATTCAAGTTTCTGAC | ^7^ |
|  | C hominis | CH LIB 13 | TCCTTGAAA TGAATATTTG TGACTCG | AAATGTGGT AGTTGCGGTT GAAA | CTTACTTCGTG GCGGCGT | ^12^ |
|  | C parvum | CP LIB13 | TCCTTGAAA TGAATATTTG TGACTCG | TTAATGTGGT AGTTGCGGTT GAAC | TATCTCTTCGT AGCGGCGTA | ^12^ |
|  | Cyclospora cayetanesis | CC 18S | AAAAGCTCGTAGTTGGATTTCTG | AACACCAACGCACGCAGC | AAGGCCGGATGACCACGA | Levy |
|  | Giardia duodenalis | 18S | GACGGCTCAGGACAACGGTT | TTGCCAGCGGTGTCCG | CCCGCGGCGGTCCCTGCTAG | ^7^ |
|  | E. histolytica | 18S | ATTGTCGTGGCATCCTAACTCA | GCGGACGGCTCATTATAACA | TCATTGAATGAATTGGCCATTT | ^7^ |
| Helminths | Ascaris | ITS1 | GTAATAGCAGTCGGCGGTTTCTT | GCCCAACATGCCACCTATTC | TTGGCGGACAATTGCATGCGAT | ^7^ |
|  | Trichuris | 18S | TTGAAACGACTTGCTCATCAACTT | CTGATTCTCCGTTAACCGTTGTC | CGATGGTACGCTACGTGCTTACCATGG | ^7^ |
|  | Necator americanus | ITS2 | CTGTTTGTCGAACGGTACTTGC | ATAACAGCGTGCACATGTTGC | CTGTACTACGCATTGTATAC | ^13^ |
|  | Ancyclostoma duodenale | ITS2 | GAATGACAGCAAACTCGTTGTTG | ATACTAGCCACTGCCGAAACGT | ATCGTTTACCGACTTTAG | ^10^ |
|  | Stongyloides stercoralis | Dispersed repetitive sequence | TCCAGAAAAGTCTTCACTCTCCAG | TGCGTTAGAATTTAGATATTATTGTTGCT | TCAGCTCCAGTTGAACAACAGCCTCCAA | ^10^ |
| Controls | 16S | Total bacteria | TCCTACGGGAGGCAGCA | GGACTACCAGGGTATCTAATCCTG | CGTATTACCGCGGCTGCT | ^7^ |
|  | CrAssphage | orf00024 | CAGAAGTACAAACTCCTAAAAAACGTAGAG | GATGACCAATAAACAAGCCATTAGC | AATAACGATTTACGTGATGTAAC | ^9^ |

| **Table S10:** Standard curve equations and efficiencies | | | | | |  |  |
| --- | --- | --- | --- | --- | --- | --- | --- |
| Pathogen | Target | Intercept | Estimate | R^2^ | Efficiency | LOQ (copies) | LOD (copies) |
| Aeromonas | aha1 | 36.39 | -3.41 | 0.99 | 97 | 10 | 10 |
| Campylobacter coli/jejuni | cadF | 36.88 | -3.53 | 0.99 | 92 | 10 | 10 |
| Campylobacter jejuni | hipO | 37.38 | -3.51 | 0.99 | 93 | 10 | 10 |
| C. coli | GlyA | 36.64 | -3.45 | 0.99 | 95 | 10 | 10 |
| E. coli/ Shigella | ipaH | 36.02 | -3.43 | 0.99 | 96 | 10 | 10 |
| EAEC | aggR | 35.54 | -3.35 | 0.96 | 99 | 10 | 10 |
| EAEC | aaiC | 35.36 | -3.43 | 0.99 | 96 | 10 | 10 |
| EAEC | aatA | 34.94 | -3.38 | 0.97 | 98 | 100 | 10 |
| Bacteroides fragilis | bft | 36.90 | -3.49 | 0.99 | 93 | 10 | 10 |
| Clostridium difficile | tcdB | 35.51 | -3.44 | 0.99 | 95 | 100 | 10 |
| EPEC | bfpA | 36.22 | -3.42 | 1.00 | 96 | 10 | 10 |
| EPEC | eae | 36.58 | -3.51 | 0.99 | 93 | 10 | 10 |
| EIEC | virF | 36.53 | -3.57 | 0.99 | 90 | 10 | 10 |
| ETEC-LT | LT | 35.84 | -3.51 | 1.00 | 93 | 10 | 10 |
| ETEC-ST | estA | 34.98 | -3.35 | 0.99 | 99 | 100 | 10 |
| EHEC 0157H7 | rfbE | 36.49 | -3.65 | 1.00 | 88 | 10 | 10 |
| Salmonella enterica | ttr | 36.56 | -3.51 | 0.99 | 93 | 10 | 10 |
| Salmonella typhi | tviB | 34.97 | -3.43 | 1.00 | 96 | 100 | 10 |
| Salmonella typhi | STY0201 | 37.78 | -3.59 | 1.00 | 90 | 10 | 10 |
| Shiga-like toxin 1 | stx1 | 36.61 | -3.46 | 0.99 | 95 | 10 | 10 |
| Shiga-like toxin 2 | stx2 | 36.11 | -3.47 | 0.99 | 94 | 10 | 10 |
| Shigella flexneri 6 | T3RE | 37.63 | -3.69 | 0.98 | 87 | 10 | 10 |
| Shigella flexneri 6 | O-antigen | 34.47 | -3.33 | 0.92 | 100 | 10 | 10 |
| Vibrio cholerae | toxR | 36.16 | -3.67 | 0.98 | 87 | 10 | 10 |
| E. coli | uidA | 37.11 | -3.67 | 0.99 | 87 | 10 | 10 |
| Helicobacter pylori | ureC | 36.51 | -3.44 | 1.00 | 95 | 100 | 10 |
| Adenovirus | Hexon | 37.62 | -3.56 | 1.00 | 91 | 10 | 10 |
| Adenovirus 40/41 | Fiber | 36.47 | -3.64 | 0.99 | 88 | 10 | 10 |
| Astrovirus | Capsid | 37.44 | -3.72 | 0.98 | 86 | 100 | 10 |
| Enterovirus | 5' UTR | 35.97 | -3.47 | 1.00 | 94 | 10 | 10 |
| Norovirus GI | ORF1-ORF2 | 36.11 | -3.60 | 0.99 | 90 | 10 | 10 |
| Norovirus GII | ORF1-ORF2 | 36.84 | -3.65 | 0.98 | 88 | 10 | 10 |
| Rotavirus A | NSP3 | 36.53 | -3.48 | 0.98 | 94 | 100 | 10 |
| Cryptosporidium spp. | 18S | 36.98 | -3.76 | 0.98 | 84 | 10 | 10 |
| C hominis | CH LIB 13 | 37.22 | -3.68 | 0.99 | 87 | 10 | 10 |
| C parvum | CP LIB13 | 35.53 | -3.50 | 0.99 | 93 | 100 | 10 |
| Cyclospora cayetanesis | CC 18S | 37.78 | -3.72 | 0.98 | 86 | 10 | 10 |
| Giardia duodenalis | 18S | 36.27 | -3.59 | 0.99 | 90 | 10 | 10 |
| E. histolytica | 18S | 36.77 | -3.43 | 0.91 | 96 | 10 | 10 |
| Ascaris | ITS1 | 36.82 | -3.50 | 0.99 | 93 | 10 | 10 |
| Trichuris | 18S | 37.48 | -3.69 | 0.99 | 87 | 10 | 10 |
| Necator americanus | ITS2 | 37.63 | -3.46 | 1.00 | 94 | 10 | 10 |
| Ancyclostoma duodenale | ITS2 | 37.41 | -3.54 | 0.99 | 92 | 10 | 10 |
| Stongyloides stercoralis | Dispersed repetitive sequence | 36.60 | -3.65 | 0.98 | 88 | 10 | 10 |

| **Table S11:** Limit of detection by sample type | |
| --- | --- |
| **Sample Type** | **LOD** |
| Feces (gene copies/g) | 370 |
| Soil (gene copies/g) | 60 |
| Drinking water (gene copies/100 mL) | 110 |
| Hand rinse (gene copies/100 mL) | 110 |

**Table S12:** gBlock sequences

**Sequence 1:**

accgctgctcattactctgatgttgatggtgagctggttggttcttcccgtctgggttggctttgaaggtaatttagatatggataatcgttatgcaccagggattagacttggttatcattttgacgatttttggcttgatcaattagaatttgggttagagcattattctgatgttaaatatacaaatactaataaaactacagatattacaagaacttatttgagtgctattaaaggtattgatgtaggtgagaaattttatttctatggtttagcagcttgcggtcatgatggacatactacttctttattgcttgctgcaaagtatttagcaagtcagaattttaatggcactttaaatctttattttcaacctgctgaagagggtttgggtggtgctaaaccaaagcttatcgtgtgcggtgcgagtgcttatgctcgtattattgacttttcaaaatttagagagattgcggatgaagttggagcttatctttttgcagacattgcacacattgctgcactccttttccgcgttccttgaccgcctttccgataccgtctctgcacgcaatacctccggattccgttttggaagcaatacatatcttagaaatgaactcatatttcttgagagaggaataaatatatcagtaagattgcaaaagaagaaatcaacagtaaatccatttatcgcaatcagattaagcagcgatacattaagacgcctaaaggatgccctgatgataatatacggaatatcaaaagtagatgcttgcagttgtccgaattgtcctcaggcatttcacgctttttcaggaattgacggtactgtttttgatttaattaatttgaagcttagggttactaaacacatacaagaccttctggagaacttttttaagagaggtgaaaaagaagttaaaatagaaattttacgtagggaatcaactaaatcaggtagtgcatactcatcatttaaggttgtttatcaggggtgtcgtctggcgaaagactgtatcattgataatttctttcagaaaagcatccagtttaattcttattctcttgatatcgaagagttagatattaataaacataacaatataaaaacgatgttaccagatataaatatagggttagggcagtatataaacaacaatcaatggttctcatctattacagacagccatttttatttatcattatcctataatcttctatcggcttatgaagcaaaagggacaaggattctaccagctttatactgggagatgagttcgcagtattacgtttttatcgcaatggcgaatccatcagctacatcgcatacaaggaagcgcaaatgatgaatgagattgccgaatggtattacctaatgctccaaatagagtatttgcttgggaaacaggatggacaccaggtttaagaagcttagaaaatgatggcacaaatggtgcttgcgcttgctgcaaccgttactgccggtgtgatgttttactaccagtctgcgtctgattccaataagtcacagaatgccatttcagaagtaatgagcgcaacgcattgatcaggatttttctggtgataatacccgtttaggtattggtggcgaatactggcgagactatttcaaaagtagcgttaacggctatttccgcatgagtccttgaaatgaatatttgtgactcgaaaaatttttagatttatttaatgtaactccagctgaattctttttcatgttctgcacaatgttcatttaactttttgtttgttttacgccgccacgaagtaagagataaattaataaattttcaaccgcaactaccacattt

**Sequence 2:**

ttcccaccggatcaccaagcttggagagaagaaccctggattcatcatgcaccacaaggttgttttaagcatgaatagtagcaattactgctgtgaattgtgttgtaatcctgcttgtaccgggtgctattaatttttttcacacttattggatggtctcaattctaactaggaccgcagaggaaagagaggaattaaggaatcaccttgcagataaactcatcgttttctcaccaggagattacaacatggctaatttaacccgtcgtcagtggctaaaagtcggtctcgccgtcggtgggatggtcacttttggtctgagcttttttgtggtaaaggaactcggtaaatatagttgtaaagtggatatttttgatccatgggtggatgccgaagaggtaagacgagagtatggcattatcccggtatcggaagtcttttcgcgaagtcagagtcgacataggcatagattttcaggccatacattaatttgccaaggttgctataaacatttgttctggagcaggctgacggaaattccgtgaactcgctggtgatcggcgttgaggtcttttttcatcgcgagttgccagaatggcatctgatgagtttccttctatgtgtccggcagatggaagagtccgtgggattacgcttttccacatcggtgtctgttattaaccacaccccaccgggcagttattttgctgtggatatacgagggcttgatgtctatcaggcgcgttttgaccttttctttcaacgcacgaatatcaacgttggggttctggatcagcaccagcttcgccagctcatccagctcgttttgcgtgactggctcgccgttgcggattttcttcagaaccgggttttgctggaacagcggctccagcgccccctgaacctgctggcgatagagtttgaaatcaatcgagcggatattagttttgcgggtctccgtctggatcaggttcttttctcctatccgtgattatagtgcatatatatattatgaattgcagtcagtttattttttaaatcaacttggcgttattttatttactttgtttttattaattaatctccttctcacgattaaaatcataaaatacagtgagttgtgtgtgcttttgtttggcgagagcaaggttttgaagtcgatgattccagcttaacccaagccatttcgactctgcgcaaaatgctcaaagattcgacaaagtccccacaatacgtcaaaacggttccgaagcgtggttaccaattgatcgcccgagtggaaacggttgaagaagagattttcggaagcaacgcgtaaactcgacccgacgcgtccgatcacctgcgtcaatgtaatgttctgcgacgctcattttgacaccagaaaaagcggctatatggtagaaaacgctttagtgagcgctttaacttccataggctataatgtgattcaaatagggcctatgcccacccctgcgattgcgtttttaaccgaagacatgcgctttttgccacggtggggtttctaaacttgttccccaggctgaagtaggtatcggtggcacgggcgaactgcaccaggcccgggctcaggtactccgaggcgtcctgcccggagatgtgcatgtaagaccactggggcttttaactttctctcttaatagacgccccacttaatgctgacacgggcactcttcgccttcaaagtgctgcacctcttggactagtggacaaaacactaaaagttttgttttctagccccct

**Sequence 3:**

cagttgcttgctgcgttcatggcagaagatcatccttttaaagtgtatgtggaacactgcctgtcacggactgctaagcagcttcgtgattctggtctcccggccagactcacagaagagcaactccatcgcatttggaggggaggaccaaagaagtgtgatggctagcaagttttccctgaatgcggctaatcctaactgcggagcacatgccctcaacccagagggtagtgtgtcgtaacgggcaactctgcagcggaaccgactactttgggtgtccgtgtttccttttattcttacattggctgcttatggtgacaatcgcttttcgctggatgcgcttccatgatctgagcatgtggacaggggatcgcgatctcctgcccgattatgtaaatgatgatggcgtctaagttttaagagccaatgttcagatggatgaggtgggagggcgatcgcaatcttgtgaatgaagatggcgtcgattttaccatcttcacgtaaccctctatgagcacaatagttaaaagctaacactgtcaaaaacctaaatggctataggggcgttatgtgaccttttgggttgtatttattagataaagaaccaatataattggtgactcataataactttacggatcactttaaatgtgacatatcattcaagtttctgacctatcagctttagacggtagggtattggccttttttctgaggaggaggtttctatcgatttgttcaaatctataaaagagatgcctttcggcaaaagaaagatctatagtttagcttgccttttatcagctgtttctttttccttgaaatgaatatttgtgactcgaaaaatttttagatttatttaatgtaactccagctgaattctttttcatgttctgcacaatgttcatttaactttttgtttgttttacgccgctacgaagagataaattaataaatgttcaaccgcaactaccacattaattttaaaagctcgtagttggatttctgtcgtggtcatccggccttgcccgtagggtgtgcgcctgggttgcccgcggcttttttccggtagccttccgcgcttcgctgcgtgcgttggtgttttttgacggctcaggacaacggttgcaccccccgcggcggtccctgctagccggacaccgctggcaattttattgtcgtggcatcctaactcacttagaatgtcatttctcaattcattgaatgaattggccattttgtactaatacaaactggatcgtctcaagtattatctttatcattcacaaagctatccttgtattttactaaccaaagaaactattactgttataatgagccgtccgcttttgtaatagcagtcggcggtttctttttttttggcggacaattgcatgcgatttgctatgtgttgagggagaataggtggcatgttgggcttttttgaaacgacttgctcatcaactttcgatggtacgctacgtgcttaccatggtgacaacggttaacggagaatcagttttctgtttgtcgaacggtacttgctctgtactacgcattgtatacgtgttcagcaattcccgtttaagtgaagaacacacgtgcaacatgtgcacgctgttatttttgaatgacagcaaactcgttgttgctgctgaatcgttcaccgactttagaacgtttcggcagtggctagtattttttccagaaaagtcttcactctccagattcagctccagttgaacaacagcctccaaaacagcaacaataatatctaaattctaacgca

| **Table S13.** Household enrollment by age group and subcounty, including number of households enrolled in longitudinal data collection. | | | | | | |
| --- | --- | --- | --- | --- | --- | --- |
|  | **Overall** | | **Turkana South** | | **Samburu North** | |
| **Age Group** | **Total** | **Longitudinal** | **Total** | **Longitudinal** | **Total** | **Longitudinal** |
| *All* | *100* | *61* | *50* | *30* | *50* | *31* |
| 0-2 months | 24 | 24 | 12 | 12 | 12 | 12 |
| 3-5 months | 22 | 21 | 11 | 10 | 11 | 11 |
| 6-11 months | 30 | 16 | 15 | 8 | 15 | 8 |
| 12-23 months | 24 | 0 | 12 | 0 | 12 | 0 |

**Table S14.** Prevalence of child diarrhea in the last 7 days, overall and by sub county.

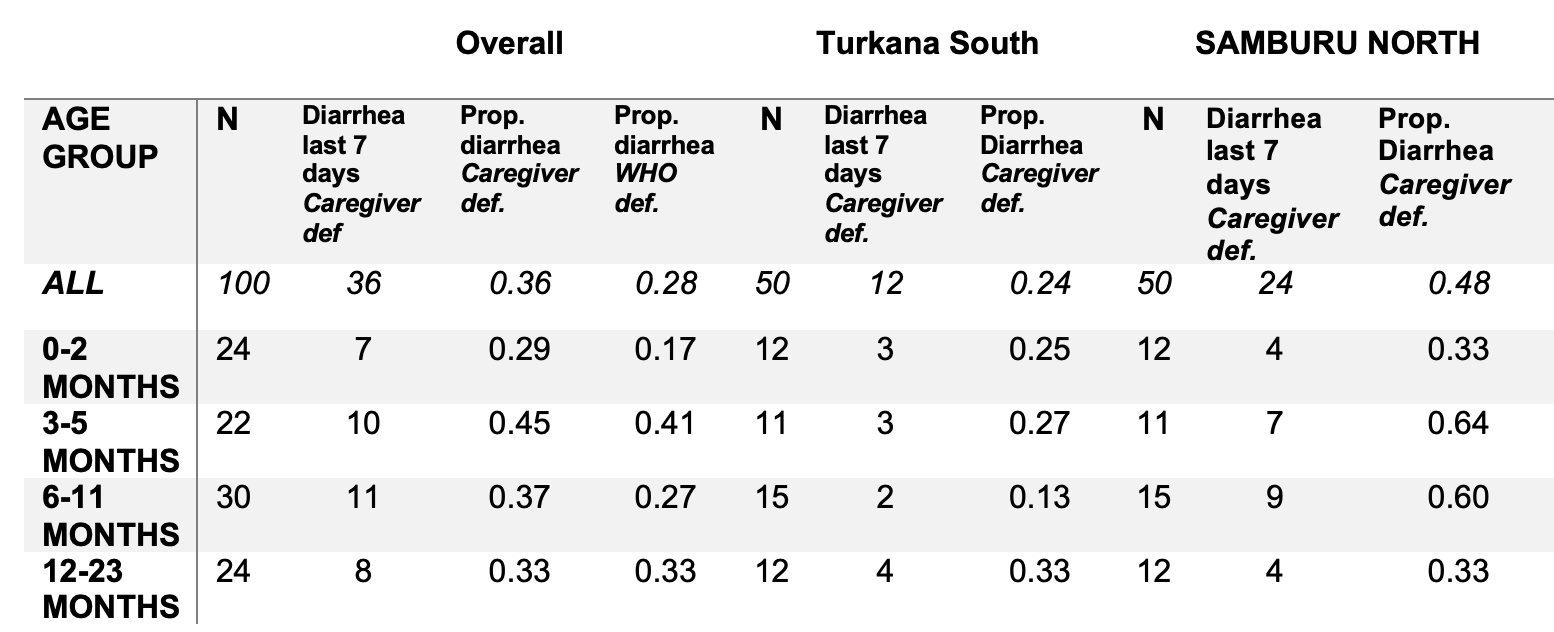

| **Table S15.** Samples collected by type. | | |
| --- | --- | --- |
| **Sample Type** | **Total** | ***E. coli* measured by culture** |
| Child Stool | 162 | 0 |
| Mother/caregiver stool | 60 | 0 |
| Animal feces | 238 | 0 |
| Soil | 163 | 73 |
| Food | 40 | 10 |
| Hand rinse | 92 | 49 |
| Drinking water (stored) | 116 | 61 |

| **Table S16**. Odds ratios of child diarrhea given pathogen detection in child stool. Bold indicates statistical significance (p < 0.05). | | |
| --- | --- | --- |
| **Pathogen** | **Odds Ratio (95% CI)** | **p** |
| Adenovirus 40/41 | 0.71 (0.29, 2.32) | 0.67 |
| Astrovirus | 2.05 (0.79, 7.36) | 0.15 |
| C hominis | 0 (0.02, 7.01) | 0.37 |
| Campylobacter coli/jejuni | 1.06 (0.42, 3.68) | 0.75 |
| Clostridium difficile | 0 (0.02, 10.66) | 0.52 |
| Cryptosporidium spp. | 1.28 (0.35, 9.81) | 0.57 |
| E. coli/ Shigella | 0.84 (0.2, 12.03) | 0.82 |
| EAEC | 0.56 (0.29, 1.21) | 0.16 |
| EHEC 0157H7 | 0 (0.02, 7.01) | 0.37 |
| EPEC | 0.62 (0.31, 1.4) | 0.28 |
| ETEC-LT | 0.6 (0.23, 2.15) | 0.50 |
| ETEC-ST | 2.61 (0.57, 34.58) | 0.21 |
| Enterovirus | 0.85 (0.45, 1.85) | 0.79 |
| Giardia duodenalis | 0.85 (0.39, 2.29) | 0.87 |
| Helicobacter pylori | 1.29 (0.42, 6.69) | 0.55 |
| Norovirus GI | 0.41 (0.11, 4.29) | 0.60 |
| Norovirus GII | 0.41 (0.15, 1.67) | 0.21 |
| **Rotavirus A** | **11.08 (1.33, 482.08)** | **0.01** |
| STEC | 1.29 (0.46, 5.39) | 0.53 |
| Salmonella enterica | 2.57 (0.31, 196.41) | 0.28 |
| Shigella flexneri 6 | 0.0 (0.03, 21.11) | 0.72 |

| **Table S17.** Results (Cq values) of inhibition testing on animal feces, human stool, and soil samples. Only targets detected within samples are displayed. | | | | | | | | | | | | | | |
| --- | --- | --- | --- | --- | --- | --- | --- | --- | --- | --- | --- | --- | --- | --- |
| **Sample** | **Goat feces** | | **Human stool** | | **Soil 1** | | **Soil 2** | | | | **Soil 3** | | | |
| **Dilution** | **1** | **1:10** | **1** | **1:10** | **1** | **1:10** | **1** | **1:10** | **1:20** | **1:50** | **1** | **1:10** | **1:20** | **1:50** |
| CADF |  |  |  |  |  |  | 32 | 36 |  |  | 46 |  |  |  |
| HIPO |  |  |  |  |  |  | 32 | 34 |  |  |  |  |  |  |
| GLYA |  |  |  |  |  |  |  |  |  |  | 35 |  |  |  |
| IPAH |  |  |  |  |  |  | 36 |  |  |  | 39 |  |  |  |
| AGGR |  |  |  |  | 38 |  | 34 |  |  |  | 34 |  |  |  |
| AAIC |  |  | 32 |  | 33 |  |  |  |  |  |  |  |  |  |
| AATA |  |  |  |  | 40 | 37 | 36 |  |  |  | 37 |  |  |  |
| BFT |  |  |  |  |  |  | 34 | 48 |  |  |  |  |  |  |
| 18s | 17 | 19 | 16 | 18 | 12 | 12 | 14 | 17 | 18 | 20 | 13 | 16 | 18 | 19 |
| BFPA |  |  | 30 | 34 |  |  |  |  |  |  |  |  |  |  |
| EAE |  |  | 32 | 35 | 33 | 35 | 29 | 32 | 33 | 35 | 36 |  |  |  |
| LT |  |  |  |  | 34 |  | 33 | 36 |  |  | 35 |  |  |  |
| ESTA |  |  |  |  |  |  | 34 |  |  |  | 30 | 35 | 32 |  |
| RFBE |  |  |  |  |  |  | 29 | 33 | 33 | 36 |  |  |  |  |
| STX1 | 33 | 36 |  |  |  |  |  |  |  |  |  |  |  |  |
| STX2 | 31 | 33 | 35 |  |  |  | 31 | 33 | 34 | 36 |  |  |  |  |
| T3RE |  |  |  |  | 40 |  |  |  |  |  |  |  |  |  |
| TOXR |  |  |  |  |  |  | 29 | 31 | 33 |  |  |  |  |  |
| UIDA | 26 | 30 | 26 | 29 | 27 | 30 | 25 | 28 | 29 | 31 | 30 | 33 | 35 |  |
| UREC |  |  | 39 |  |  |  |  |  |  |  |  |  |  |  |
| HEXON |  |  |  |  |  |  | 35 |  |  |  |  |  |  |  |
| 5_UTR |  |  |  |  |  |  | 32 | 38 | 44 |  |  |  |  |  |
| NGI_ORF1-ORF2 |  |  |  |  |  |  | 35 |  | 36 |  |  |  |  |  |
| NSP3 |  |  |  |  |  |  | 33 | 36 | 34 |  |  |  |  |  |
| CP_18S | 35 | 40 |  |  | 22 | 21 | 26 | 28 | 29 | 32 | 28 | 31 | 32 | 34 |
| CC_18S |  | 38 |  |  | 36 | 33 | 34 | 35 | 36 | 40 | 41 | 40 |  |  |
| G_18S |  |  |  |  | 29 | 29 | 30 | 35 | 32 |  | 30 | 33 |  |  |
| synthetic construct | 33 |  | 33 | 36 | 34 |  | 32 |  | 36 |  | 33 | 35 | 36 |  |
| ORF00024 | 8 | 11 | 5 | 7 | 5 | 5 | 5 | 8 | 9 | 12 | 5 | 10 | 12 | 14 |

### Figures

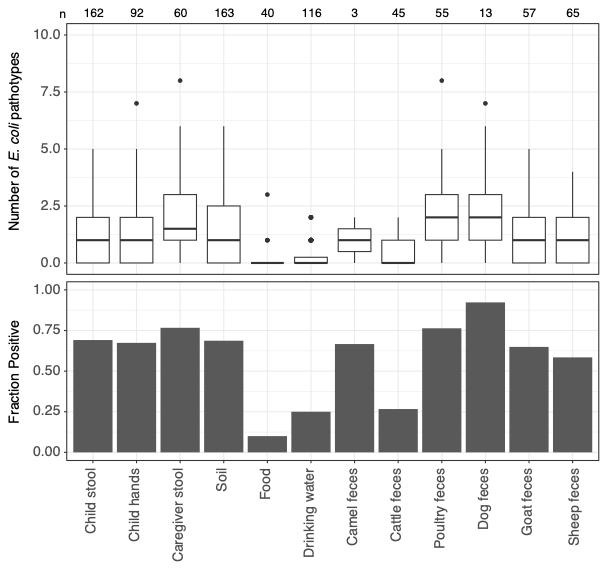

**Figure S1.** *E. coli* TAC detections by pathotype and sample. Top shows a boxplot of the number of pathotypes present in each sample. Bottom displays the fraction of samples positive for any *E. coli* pathotype by sample type.

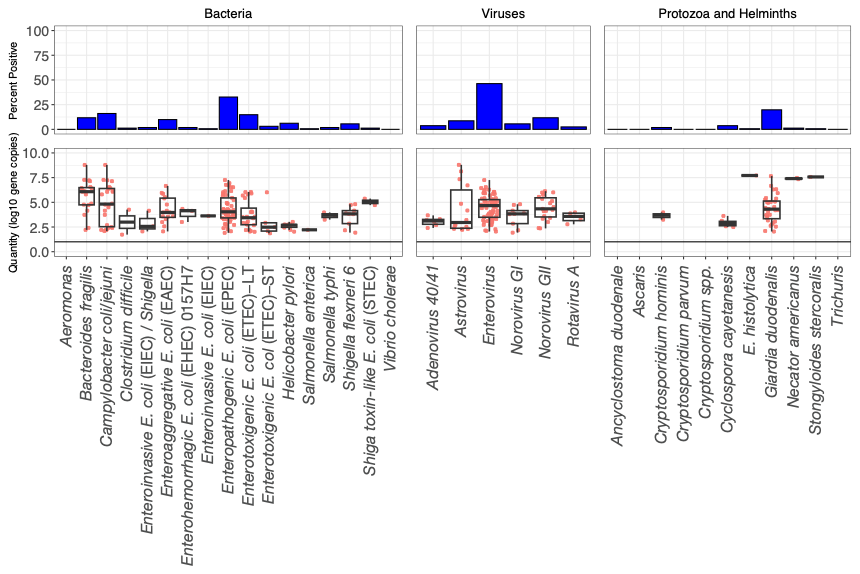

**Figure S2:** Pathogen positivity rates and quantities in child stool by pathogen, n = 162.

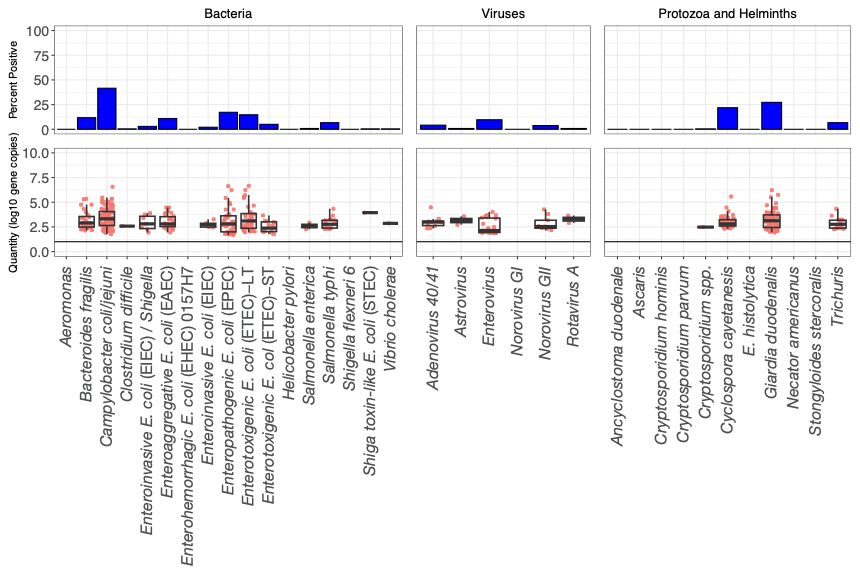

**Figure S3:** Pathogen positivity rates and quantities in animal feces by pathogen, n = 238.

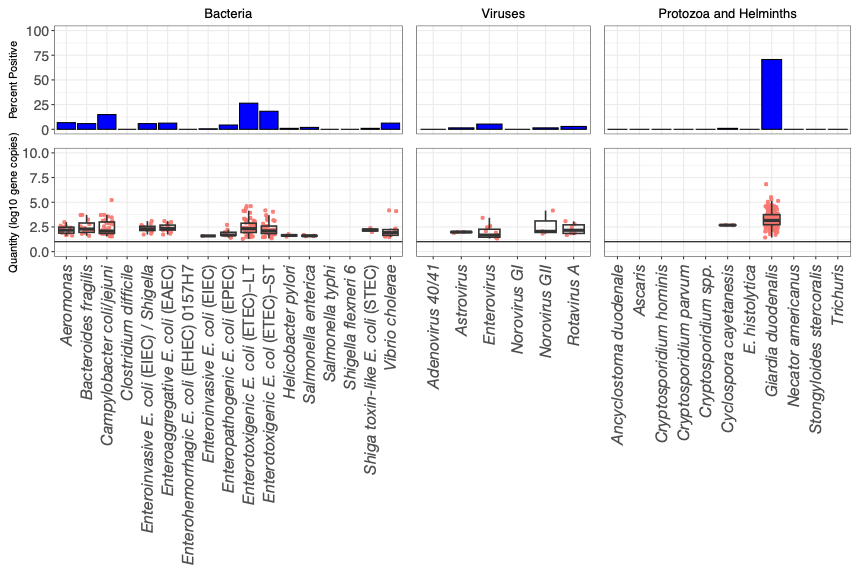

**Figure S4:** Pathogen positivity rates and quantities in drinking water and child hand rinse samples by pathogen, n = 208.

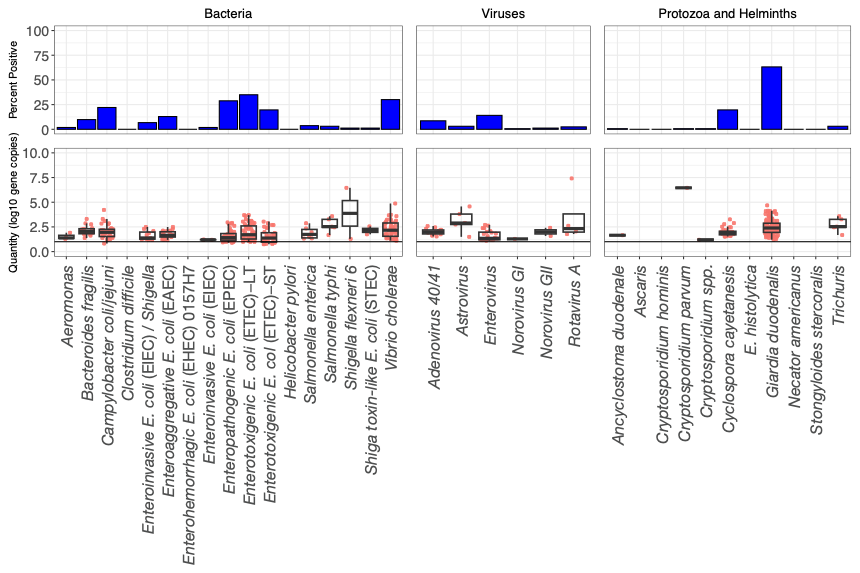

**Figure S5:** Pathogen positivity rates and quantities in soil by pathogen, n = 163.

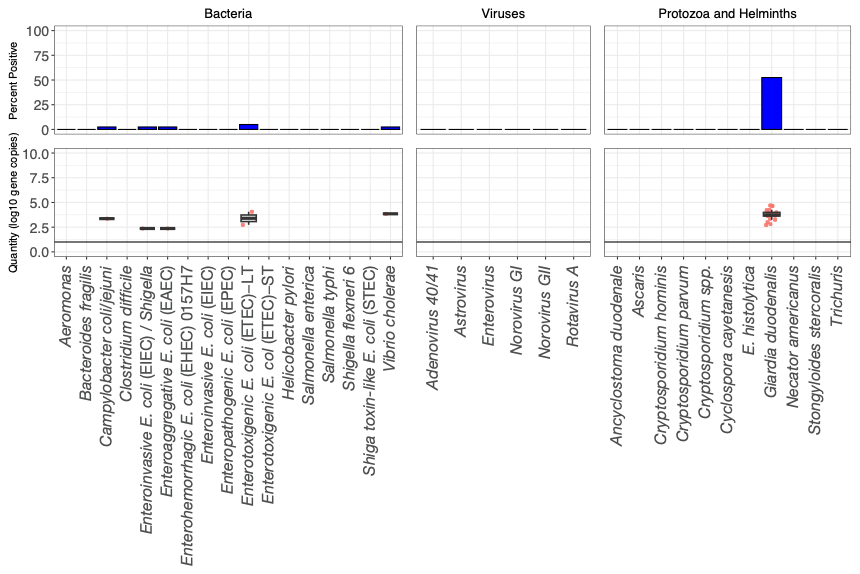

**Figure S6:** Pathogen positivity rates and quantities in food by pathogen, n = 40.

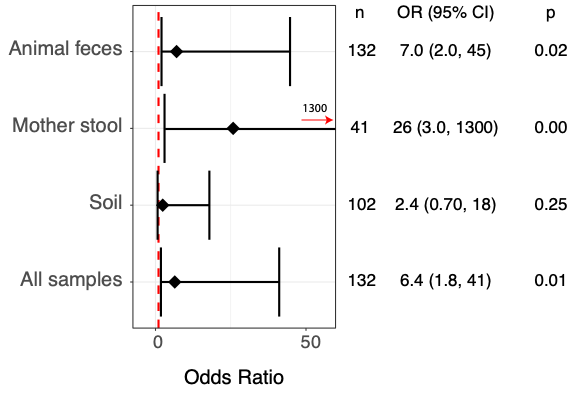

**Figure S7:** Odds ratios of pathogen detection in animal feces or environmental samples and subsequent or cooccurring new infection (both are included) with the same pathogen in children. Bars represent 95% confidence intervals. The dotted red line indicates an odds ratio of 1. The environmental samples category includes hands, soil, water, and food. Food alone is not displayed because of uncertainty around the point estimate.

**Figure S8. Pathogen Transmission Networks**

Graphs of pathogen transmission networks for pathogen detection lagged one visit in blue, and co-detection in the same sampling visit to a household in red; the arrow direction indicates the sample type associated with detection in the other sample type on the prior visit. Nodes are sample types; edges are the odds ratios of pathogen co-occurrence between pairs. Only lines for odds ratios with a p-value < 0.05 are presented. Edge weights correspond to the value of the odds ratio; a thicker edge indicates greater odds of co-occurrence.

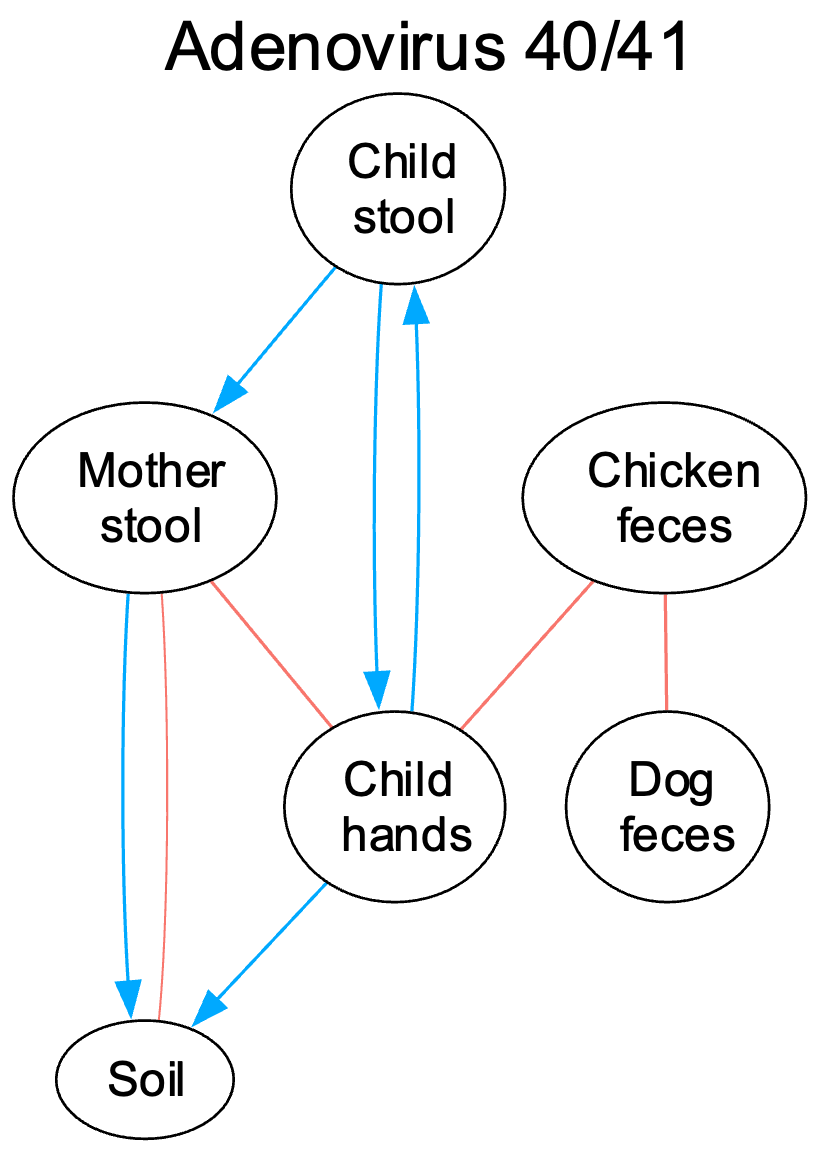

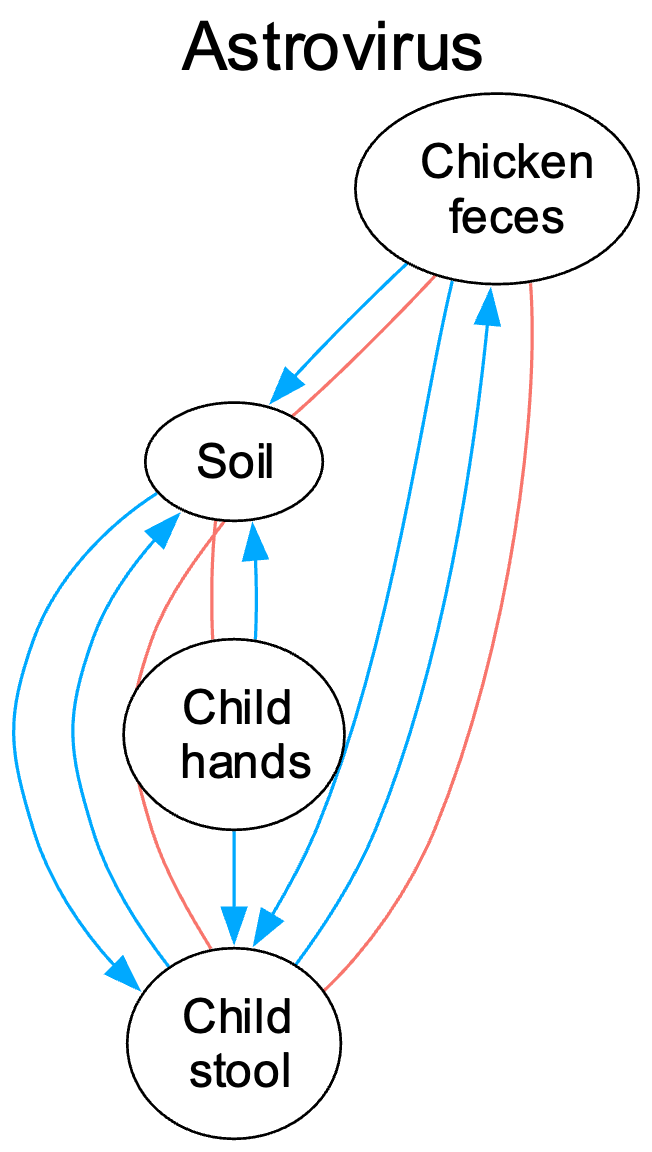

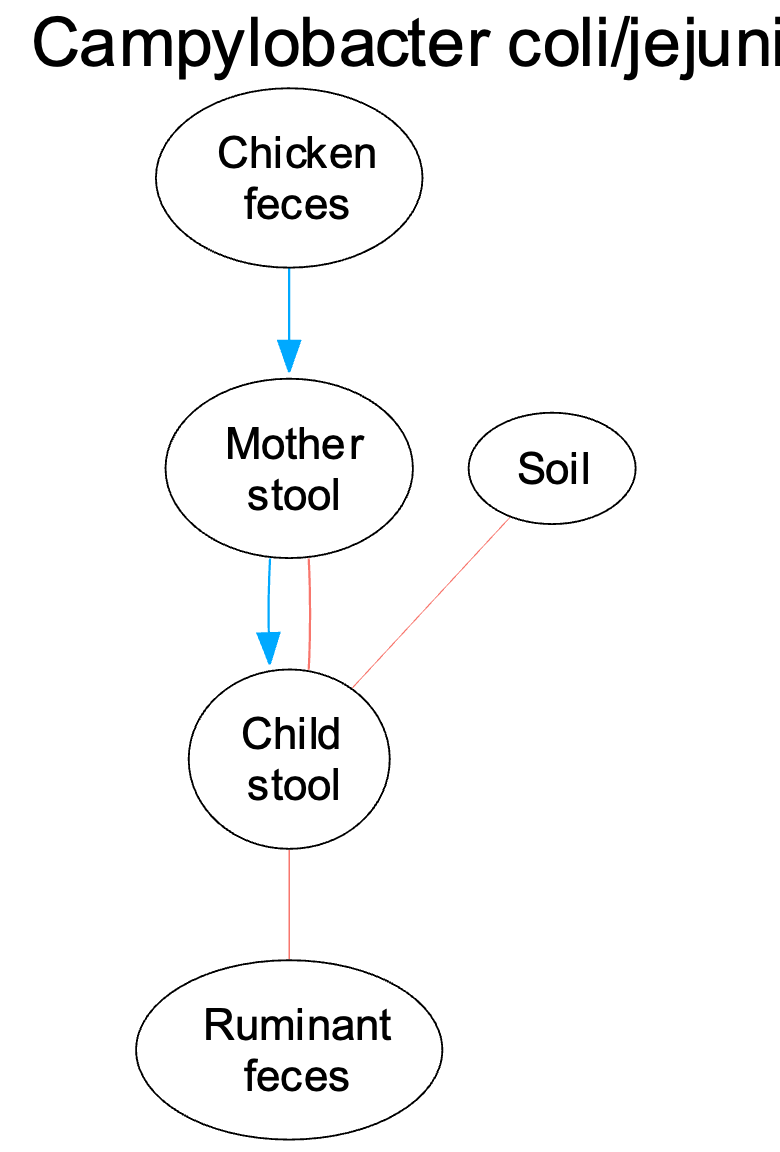

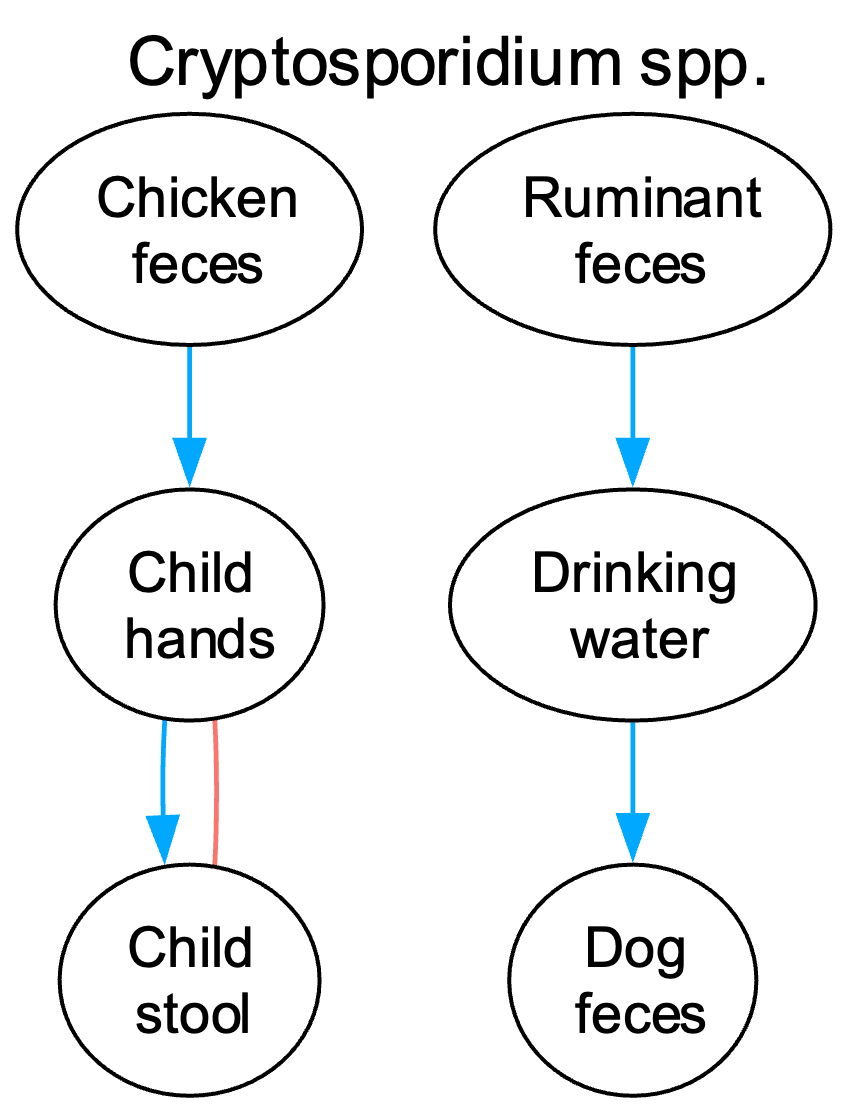

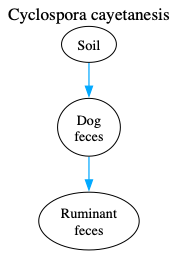

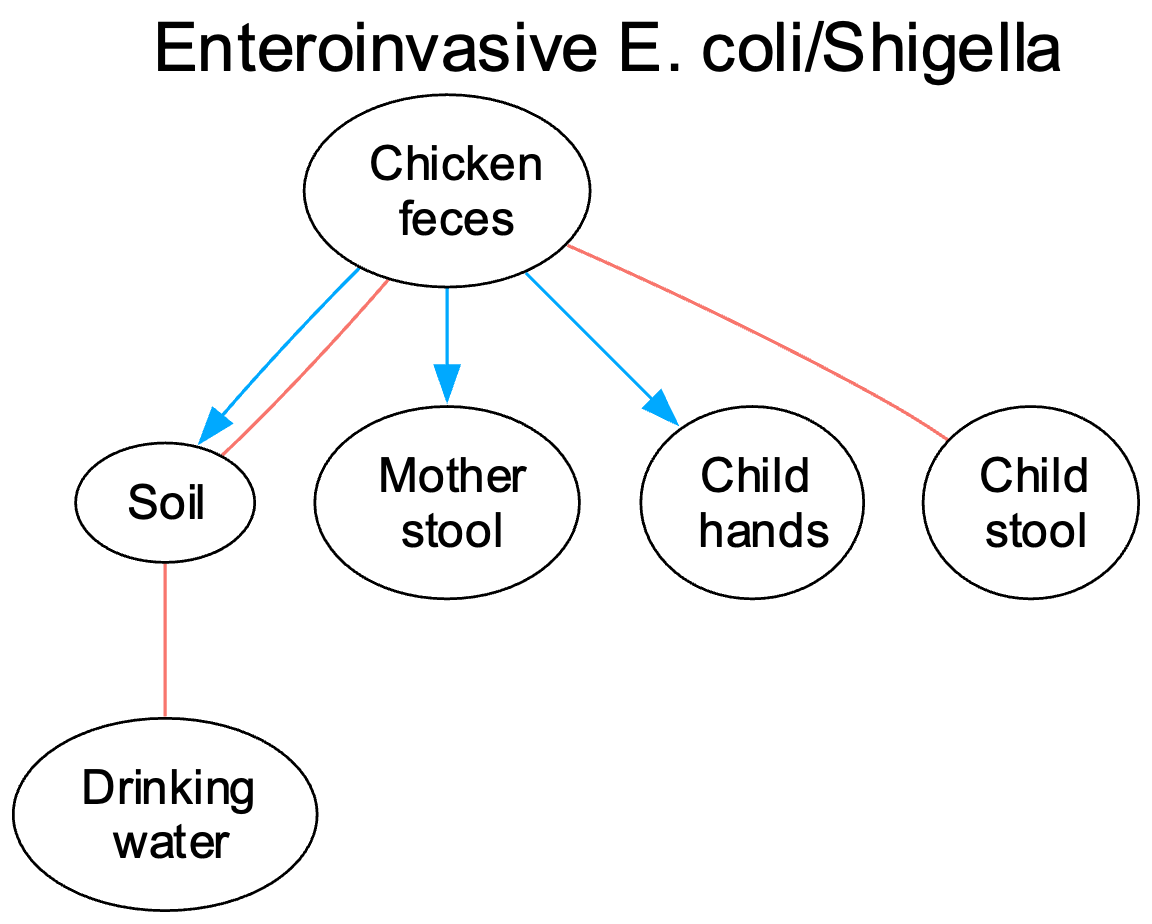

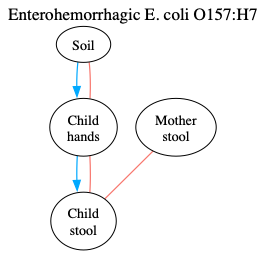

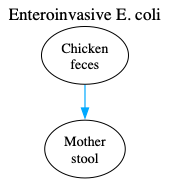

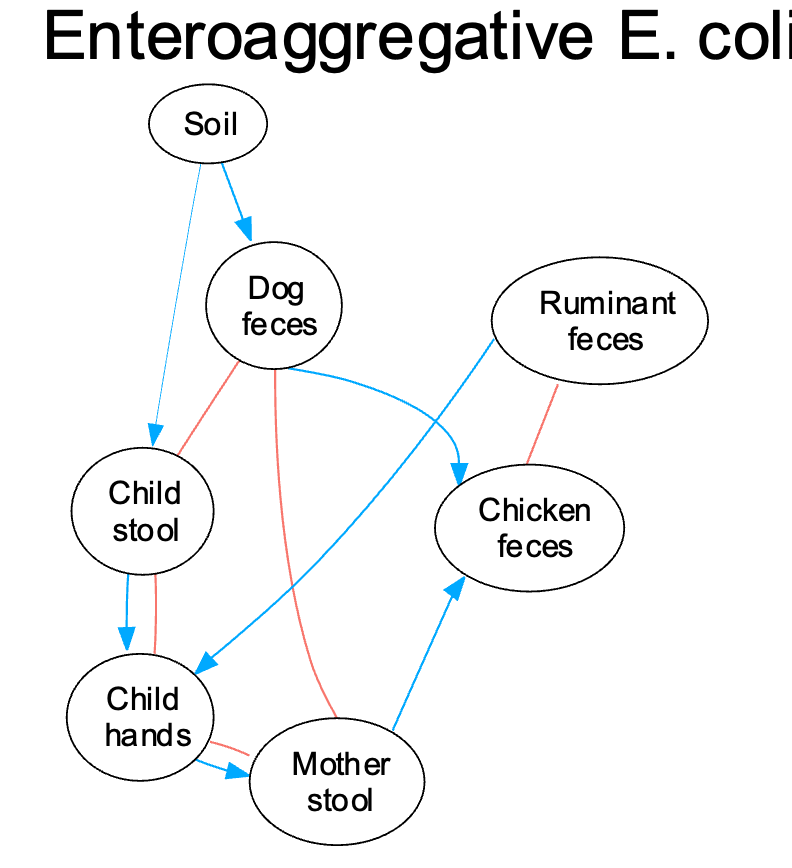

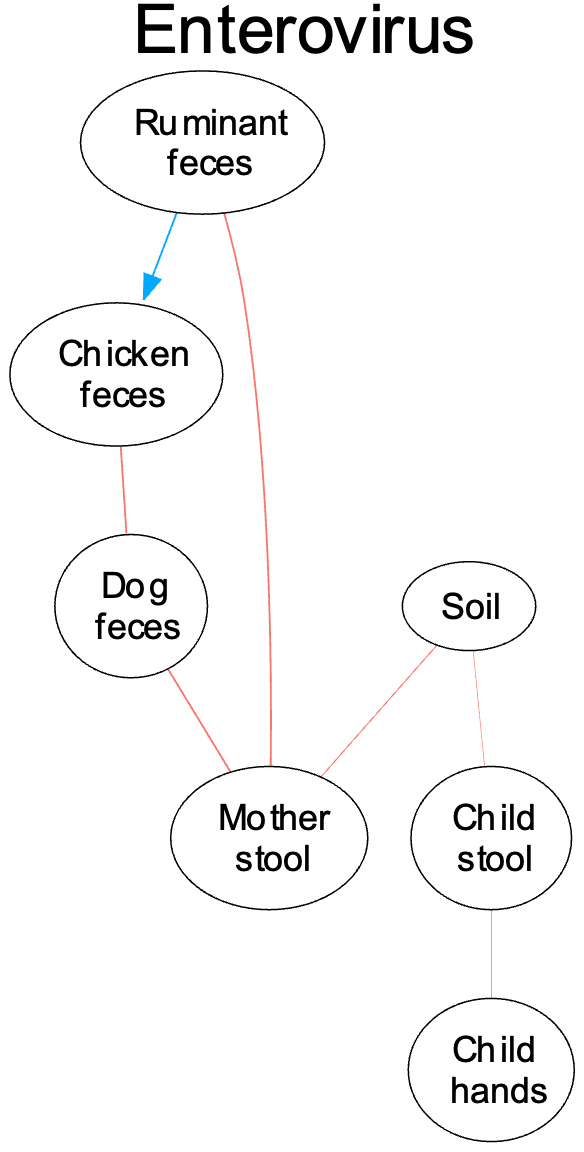

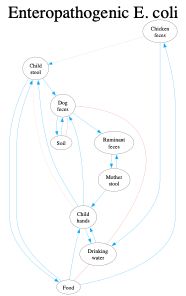

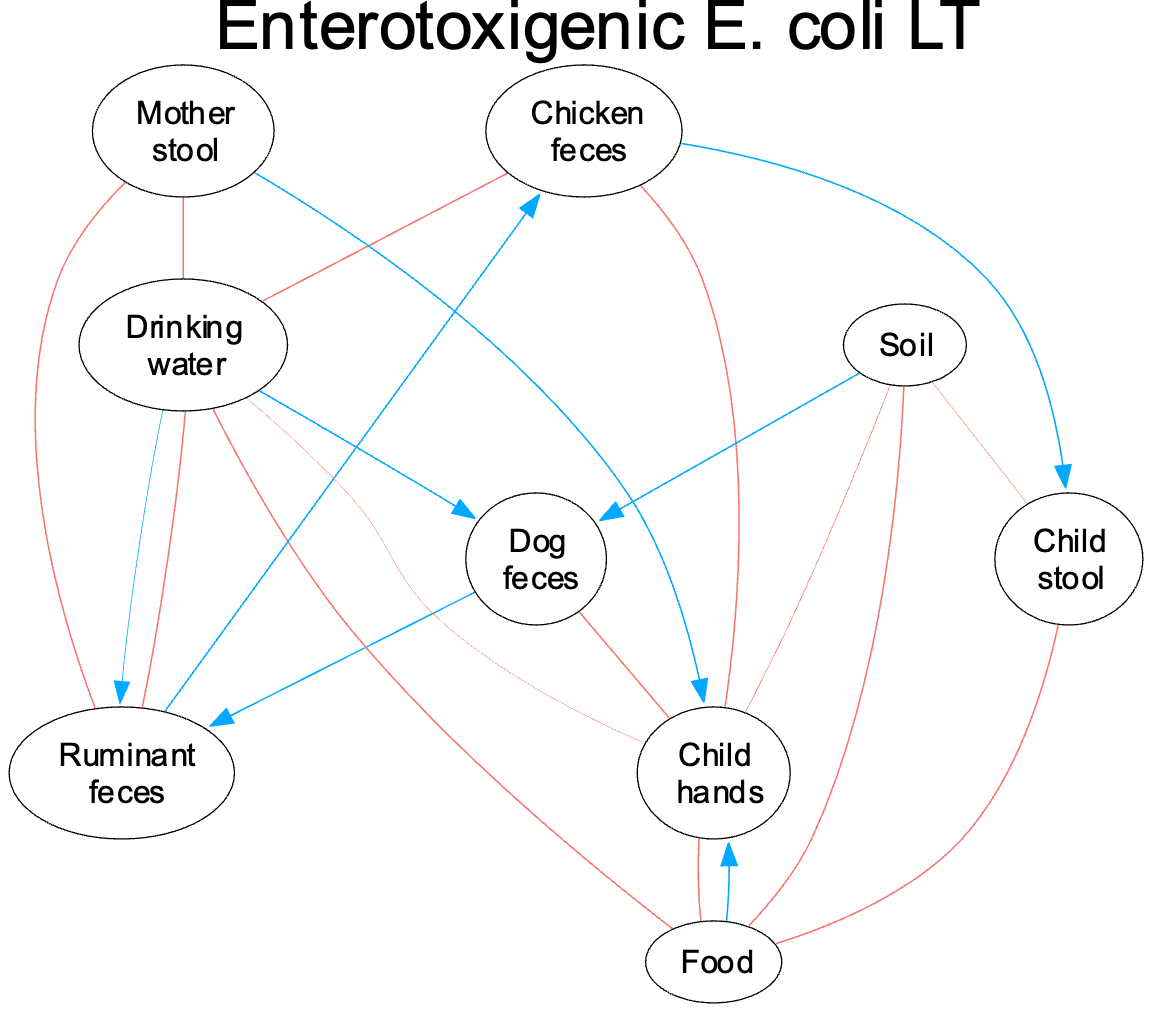

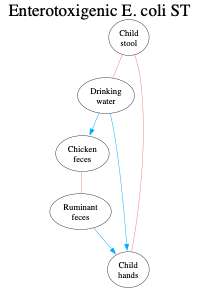

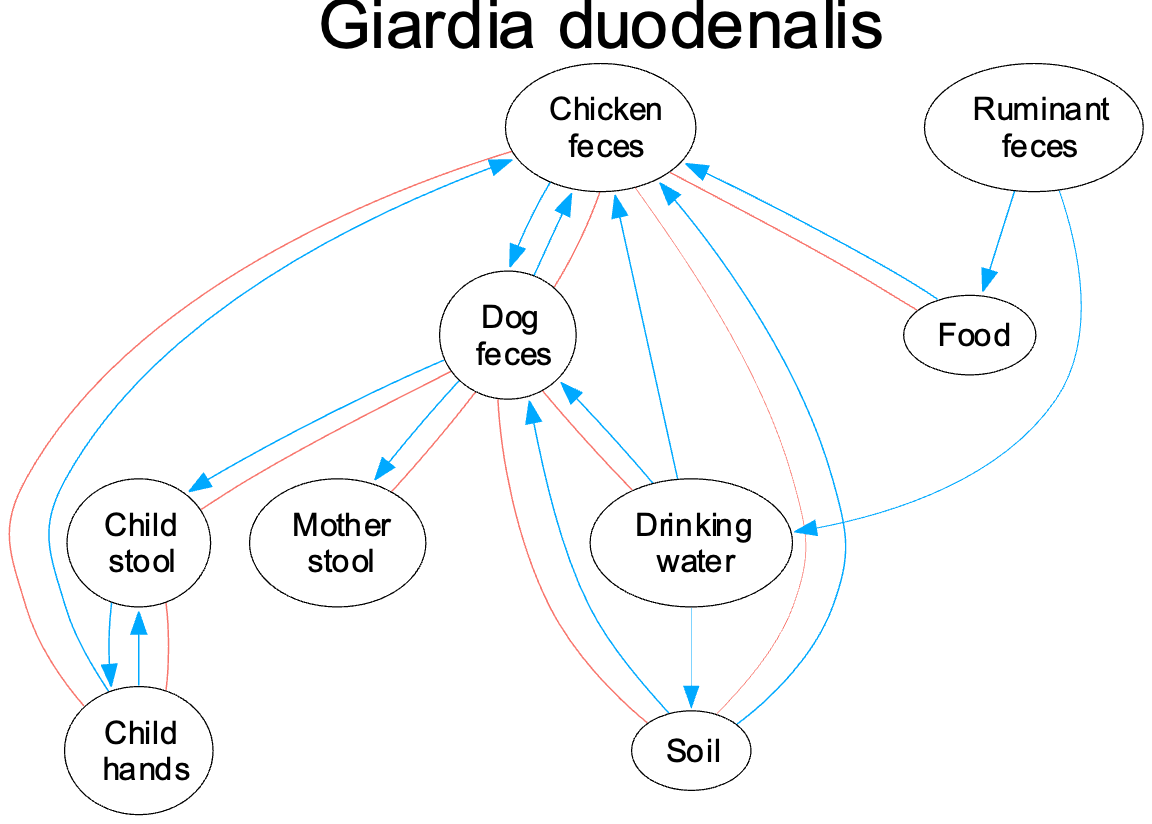

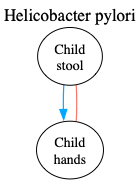

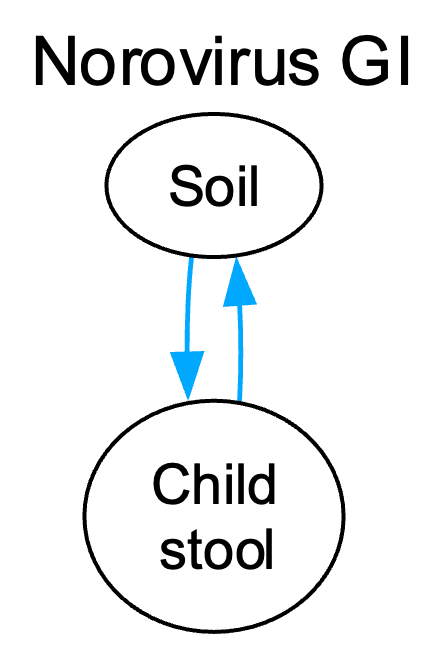

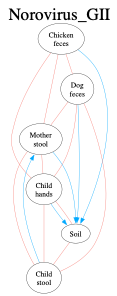

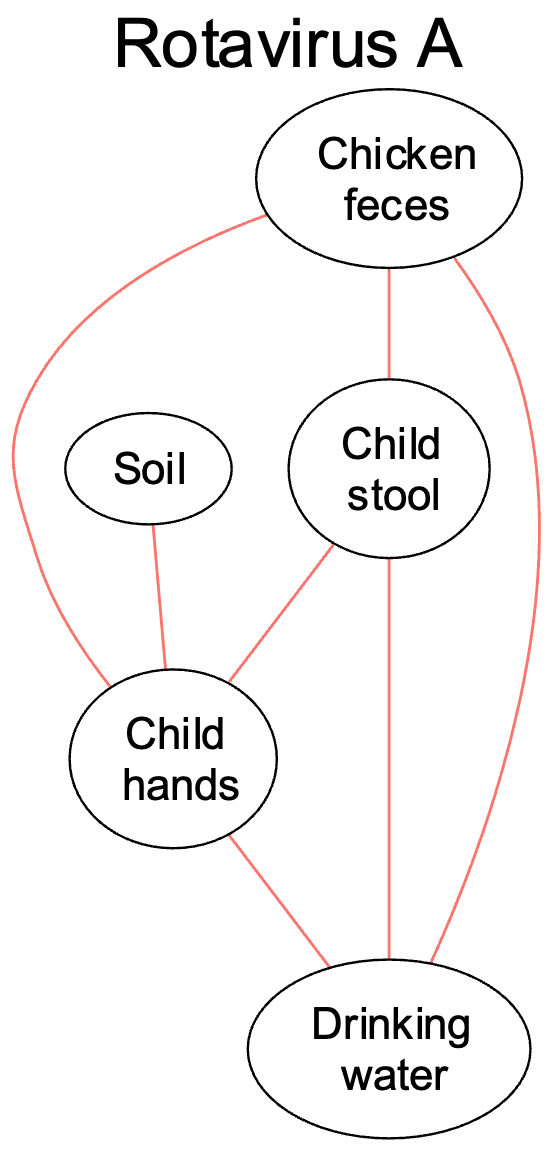

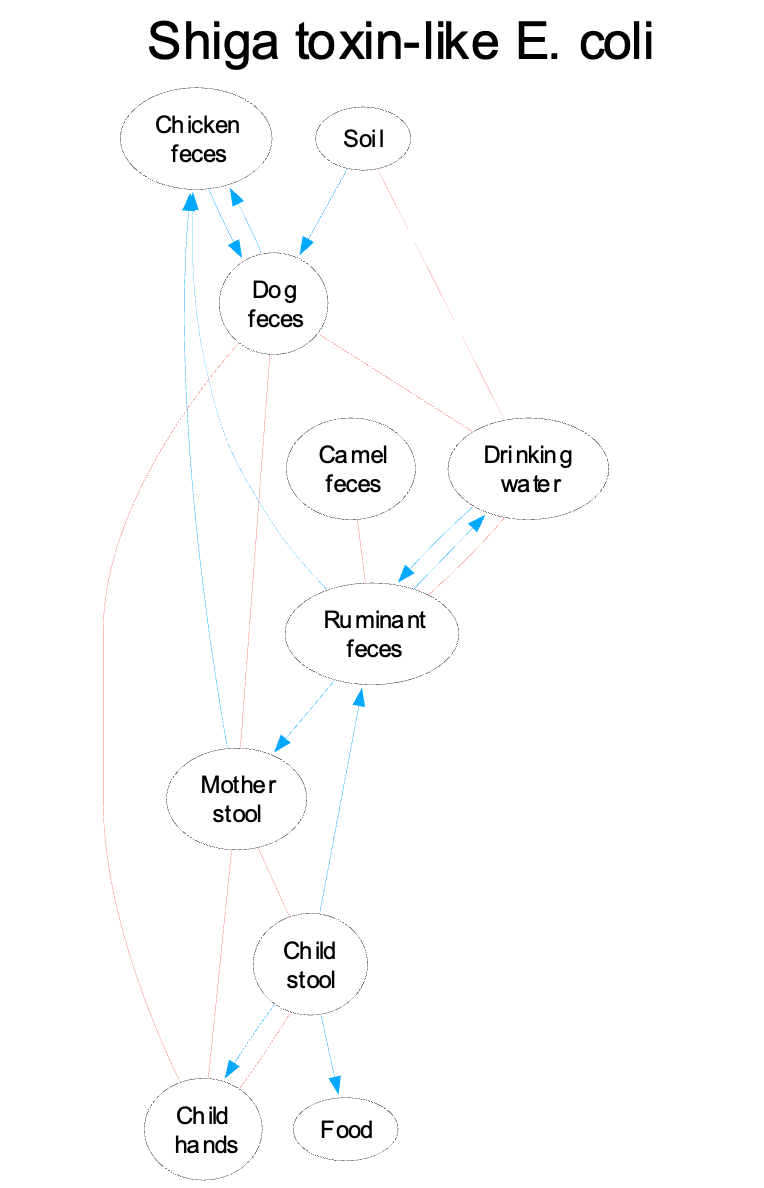

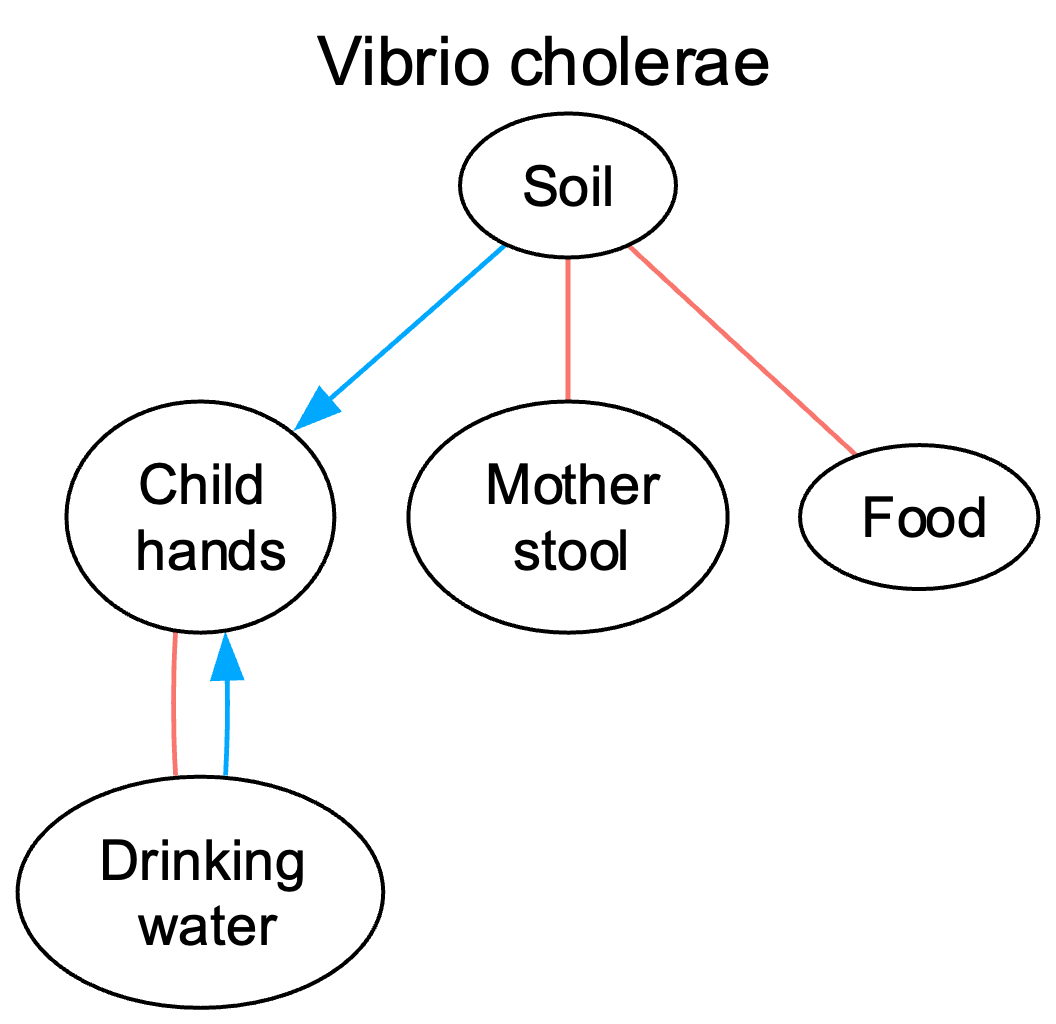

**References**

1.	IPA & Poverty Probability Index. *Kenya 2015 PPI*. (2018).

2.	Sulis, G. *et al.* Antibiotic prescription practices in primary care in low- and middle-income countries: A systematic review and meta-analysis. *PLOS Med.* 17, e1003139 (2020).

3.	Miller, J. E. *et al.* Maternal antibiotic exposure during pregnancy and hospitalization with infection in offspring: a population-based cohort study. *Int. J. Epidemiol.* 47, 561–571 (2018).
